# Single-Nucleus Atlas of Cell-Type Specific Genetic Regulation in the Human Brain

**DOI:** 10.1101/2024.11.02.24316590

**Authors:** Biao Zeng, Hui Yang, N.M. Prashant, Sanan Venkatesh, Deepika Mathur, Pavan Auluck, David A. Bennett, Stefano Marenco, Vahram Haroutunian, PsychAD Consortium, Georgios Voloudakis, Donghoon Lee, John F. Fullard, Jaroslav Bendl, Kiran Girdhar, Gabriel E. Hoffman, Panos Roussos

## Abstract

Genetic risk variants for common diseases are predominantly located in non-coding regulatory regions and modulate gene expression. Although bulk tissue studies have elucidated shared mechanisms of regulatory and disease-associated genetics, the cellular specificity of these mechanisms remains largely unexplored. This study presents a comprehensive single-nucleus multi-ancestry atlas of genetic regulation of gene expression in the human prefrontal cortex, comprising 5.6 million nuclei from 1,384 donors of diverse ancestries. Through multi-resolution analyses spanning eight major cell classes and 27 subclasses, we identify genetic regulation for 14,258 genes, with 857 showing cell type-specific regulatory effects at the class level and 981 at the subclass level. Colocalization of genetic variants associated with gene regulation and disease traits uncovers novel cell type-specific genes implicated in Alzheimer’s disease, schizophrenia, and other disorders, which were not detectable in bulk tissue analyses. Analysis of dynamic genetic regulation at the single nucleus level identifies 2,073 genes with regulatory effects that vary across developmental trajectories, inferred from a broad age range of donors. We also uncover 1,655 genes with *trans*-regulatory effects, revealing distal regulation of gene expression. This high-resolution atlas provides unprecedented insight into the cell type-specific regulatory architecture of the human brain, and offers novel mechanistic targets for understanding the genetic basis of neuropsychiatric and neurodegenerative diseases.

## Introduction

The human brain is composed of an array of cell types with a range of biological functions, cellular interactions and unique contributions to the genetic risk for neurodegenerative and neuropsychiatric traits ^1–3^. Genetic risk variants identified by large-scale genome-wide association studies (GWAS) are predominantly located in non-coding regions and play a regulatory role in modifying gene expression ^4–9^. Understanding the role of shared regulatory and disease risk effects in bulk tissue has yielded novel insight into the genes and molecular mechanisms underlying disease biology ^7,10–12^. Yet analysis of bulk tissue cannot examine genetic regulatory effects that vary across the array of cell types, and overlooks the distinct role of these cell types in disease biology ^13–15^. Recent efforts to increase the cell type resolution of genetic regulatory atlases using cell sorting or gene expression imputation based on single cell reference panels have offered some improvement, especially for common cell types ^8,9,16–18^

Advances in single-cell and -nucleus RNA-seq have enabled the collection of transcriptional profiles of diverse cell types and offer an unbiased strategy to study genetic regulatory variants affecting gene expression in each cell type ^19,20^. Recent analyses of genetic regulation in single nucleus transcriptome data from postmortem human brain identified genetic regulatory effects for broad cell categories and identified genes, cell types and molecular processes implicated in the biology of brain-related traits ^21–23^. Yet constructing a genetic regulatory atlas with a larger sample size, genetic diversity, number of nuclei and RNA-seq reads can increase statistical power, cellular resolution and coverage of rare cell types with key roles in disease biology.

In this work, we collected 5.6 million nuclei from the human dorsolateral prefrontal cortex of a genetically diverse set of 1384 donors from the full PsychAD dataset, of which 35.6% are of non-European ancestry ^24^. We performed genetic regulatory analyses at two cellular resolutions with nuclei annotated into 8 cell classes and 27 subclasses. Integrating our regulatory atlas with disease risk variants using colocalization analyses identifies cell types and genes in disease biology of brain-related traits. Analysis of *trans*-regulatory effects and dynamic regulatory effects that change over the course of a developmental trajectory offer further insight into the complexity of the genetic architecture of gene expression. This multi-resolution genetic regulatory atlas of gene expression in the human brain improves our understanding of the molecular mechanisms affecting gene expression and disease risk.

## Results

### Genetic regulation of gene expression in the human brain

To characterize the genetic regulation of gene expression across cell types in the human brain, we obtained postmortem tissue specimens from the dorsolateral prefrontal cortex (DLPFC) from 3 brain banks to create a genetically diverse set of 1,384 donors with genotype data from the full PsychAD dataset ^24,25^ of which 493 (35.6%) are of non-European ancestry (**Fig 1A**). Single nucleus RNA sequencing (snRNA-seq) was performed on postmortem tissue yielding 5.6M nuclei after quality control, and nuclei were annotated to 8 cell classes and 27 subclasses (**Fig 1B**). Analysis of genetic variants within 1Mb of the transcription start site identified expression quantitative trait loci (eQTL) at the class and subclass level (**Fig 1C**). Genome-wide, eQTL results were highly concordant with results from other single nucleus data and had the highest concordance for matching cell types ^21^ (**Fig S1**). The number of genes with statistically significant eQTLs (i.e. eGenes) varied widely with 10,913 eGenes detected in excitatory neurons (EN), but only 414 in endothelial cells at the class level. Similarly, at the subclass level, 8,812 eGenes were detected in layer 2/3 intratelencephalic (IT) excitatory neurons (EN_L2_3_IT) but only 1,683 in layer 6b excitatory neurons (EN_L6B). Analysis at the subclass level increases the cellular resolution, while sacrificing some statistical power to detect effects shared across many subclasses (i.e. excitatory neurons). Beyond differences in the genetic regulatory landscape across cell types, statistical power to detect genetic regulatory effects is heavily influenced by other factors. Indeed, the number of detected eGenes increases with cell type abundance (**Fig 1D**) and the average read count per cell type (**Fig S2**). Furthermore, we evaluated the concordance of genetic effect sizes at the class- and subclass level compared to bulk-level analysis aggregating all nuclei. Concordance increased substantially with cell type abundance, with neuronal classes and subclasses showing markedly higher concordance than non-neurons (**Fig 1E**), consistent with the higher RNA production in neurons (**Fig S2**).

**Figure 1.**
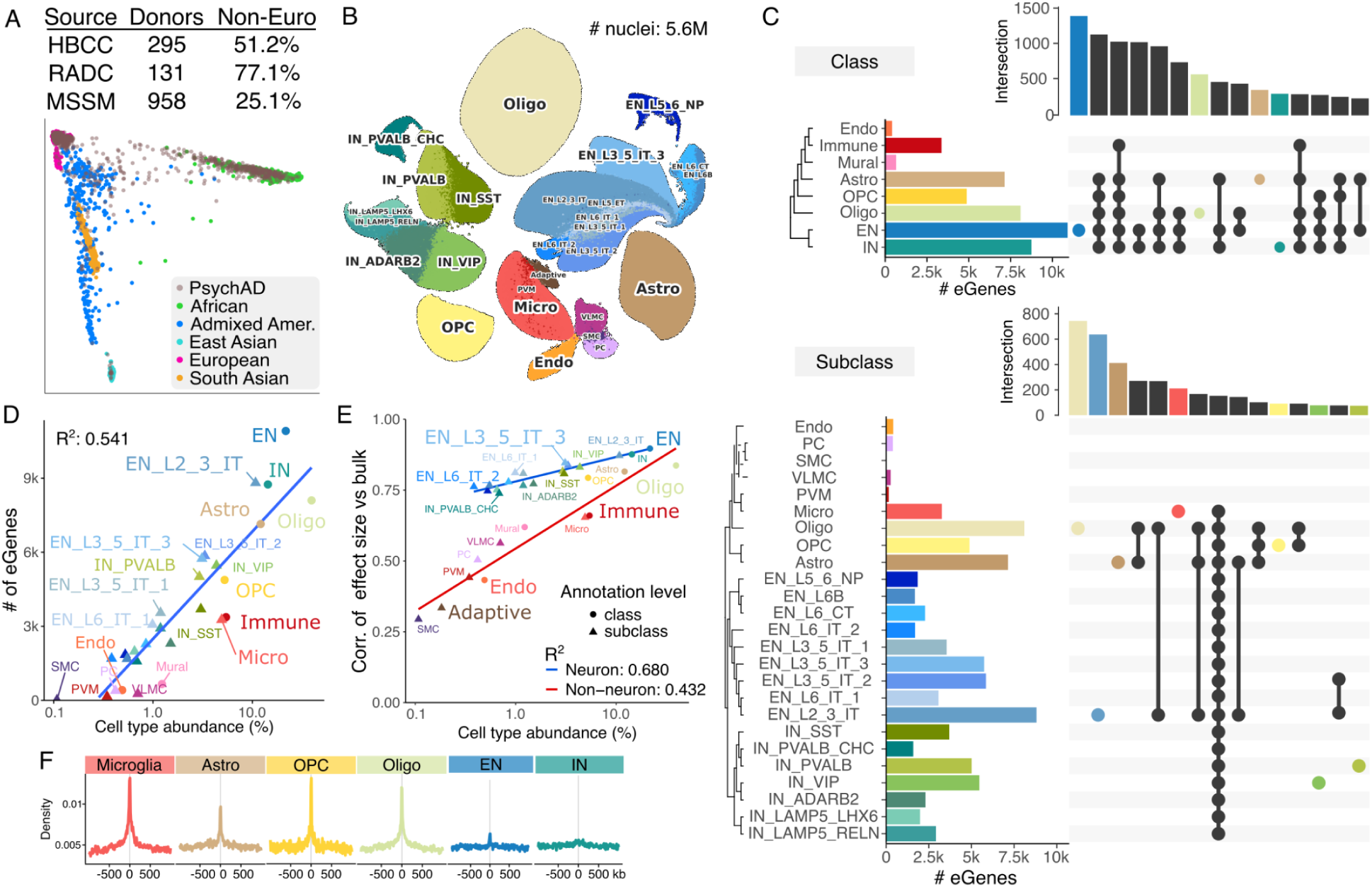
Genetic regulation of gene expression at the cell-type level. **A)** Genetic ancestry of 1,384 donors with genotype data by brain bank. Tissue specimens were obtained from the Human Brain Collection Core (HBCC), Rush Alzheimer’s Disease Center (RADC), and Mount Sinai School of Medicine (MSSM). Principal components analysis of donors from the PsychAD cohort and 1000 Genomes Project show diverse genetic ancestry (bottom). **B)** UMAP plot of 5.6M nuclei annotated for 27 cell subclasses. **C)** Upset plot for the number of eGenes detected for each class (top) and subclass (bottom) along with counts of eGenes detected in multiple classes or subclasses. **D)** The number of eGenes detected in each class or subclass shown as a function of cell type abundance on a log_10_ scale. Blue line indicates least squares fit. Squared Pearson correlation is shown. **E)** For each class or subclass, the correlation in estimated genetic effect sizes from bulk-level analysis aggregating all nuclei as a function of cell type abundance on a log_10_ scale. Least square fits are shown for neurons (blue) and non-neurons (red). Annotation level is indicated by a circle for class and a triangle for subclass. **F)** Enrichment of lead eQTL variants near open chromatin regions from six matching cell-type populations from single-cell ATAC-seq data ^26^.

Regulatory variants identified in each cell class captured biology specific to that lineage. Lead eQTL variants were enriched around open chromatin regions identified from the corresponding cell types ^26^ (**Fig 1F**, **S3**). This is consistent with cell type-specific regulatory programs especially for glia, with lower enrichment for neurons as seen previously ^9,21^. Integrating with a massively parallel reporter assay (MPRA) in human induced pluripotent stem cell-derived NGN2 excitatory neurons ^9^, indicated allelic effect size in the experiment was best predicted by fine-mapped regulatory variants in excitatory neurons (**Fig S4**).

### Cell-Type and Trait-Specific Insights into Neuropsychiatric and Neurodegenerative Disorders

Integrating this catalog of genetic regulatory variation with genetic risk for complex traits can identify genes and cell types underlying disease biology. Pairs of cell types and traits where regulatory variants from statistical fine-mapping are enriched for trait heritability were identified using stratified LD score regression after accounting for baseline annotations ^3^ (**Fig 2A, S5**). Neuronal regulatory variants are enriched for heritability for neuropsychiatric traits with schizophrenia (SZ) showing the broadest enrichment followed by bipolar disorder (BD) and major depressive disorder (MDD). Yet these traits also show enrichment in astrocytes and oligodendrocytes, and SZ and MDD also show enrichment in OPCs. The neurodegenerative traits Alzheimer’s disease (AD) and Parkinson’s disease (PD) show enrichment in microglia but not neuronal subclasses. In analysis at the class level, where there is increased power to detect effects shared across many subclasses, PD, multiple sclerosis (MS) and amyotrophic lateral sclerosis (ALS) show enrichment in neurons (**Fig S5**). Other complex traits examined do not show enrichment for brain regulatory variants, with the exceptions of type 2 diabetes and body mass index which are metabolic traits with a behavioral component. In addition, heritability mediation analysis ^27^ finds that regulatory variants in specific cell types also mediate a significant fraction of the heritability for complex traits, with notable signals for SZ in neurons and AD in microglia (**Fig S6**).

**Figure 2:**
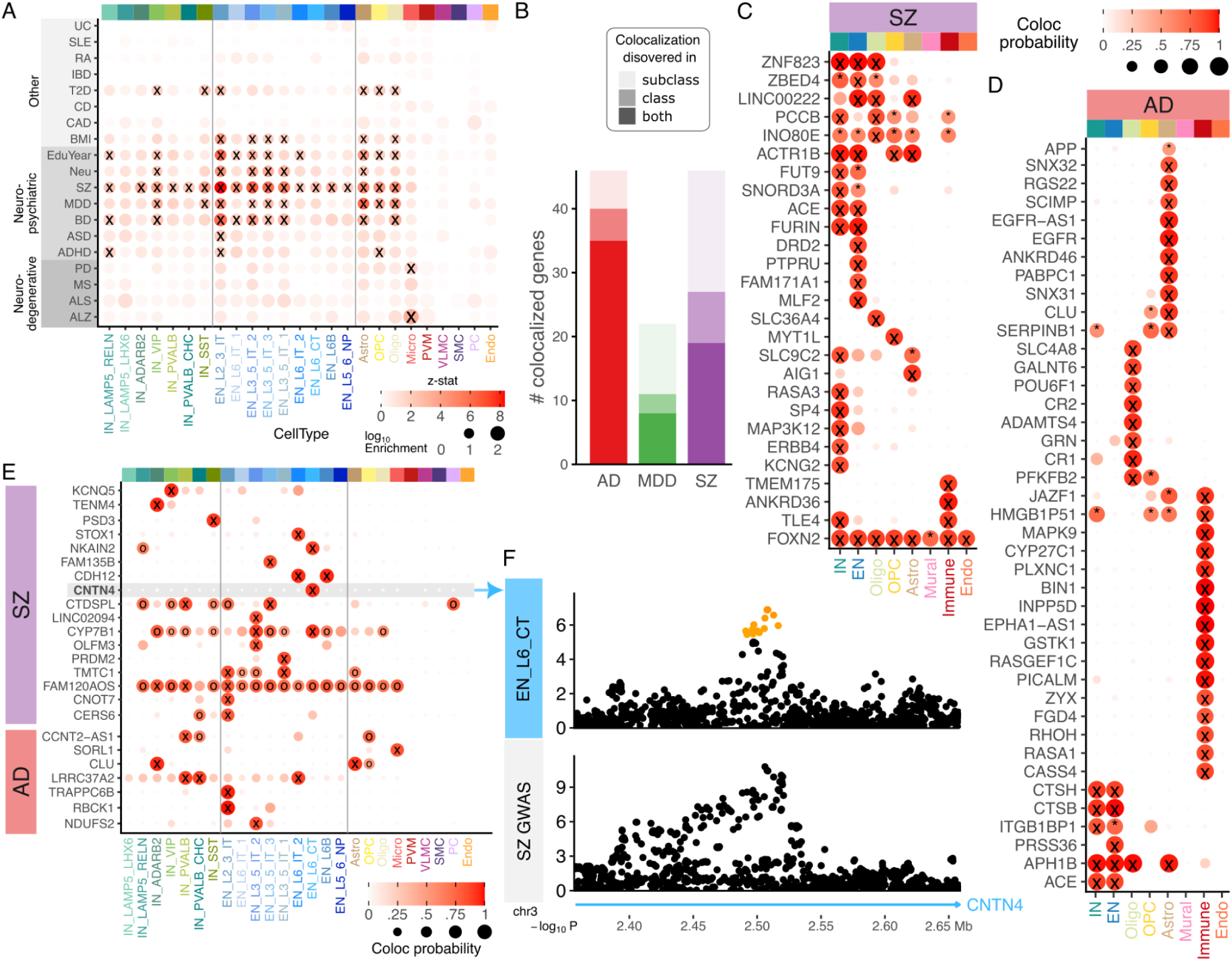
Integrative analysis of variants associated with gene expression and disease risk. **A)** Enrichment of genetic regulatory variants in the 95% credible set from statistical fine-mapping for heritability of genetic traits. Color indicates the z-statistic of the null hypothesis of no enrichment, point size indicates log10 enrichment. Tests with FDR < 5% are indicated by ‘X’. **B**) Number of genes where signals for genetic regulation of gene expression and disease risk colocalize with posterior probability > 0.8. Numbers are shown for cell type class, subclass and intersection. **C,D**) Colocalization signal at the class level for **C**) SCZ and **D**) AD. Posterior probabilities > 0.8 are indicated by ‘X’, and > 0.5 are indicated by ‘*’. **E**) Colocalization signals at the subclass level for SCZ and AD that are not found at the class level. Posterior probabilities > 0.8 are indicated by ‘X’, and > 0.5 are indicated by ‘o’. **F**) Manhattan plot of colocalization between regulation of CNTN4 expression in astrocytes and genetic risk for AD. Color indicates posterior inclusion probability from statistical fine-mapping.

Colocalization analysis of genetic signals shared between regulatory and risk variants identified genes involved in the molecular etiology of disease, including 46 genes in AD, 22 in MDD and 46 in SZ (**Fig 2B**). While some genes are shared across class and subclass-level analyses, many are only identified at one level, highlighting the importance of multi-resolution analyses. At the class-level many of the colocalized genes for SZ are identified in only excitatory and inhibitory neurons such as FUT9, SNORD3A, ACE, FURIN, while others like ACTR1B and ZNF832 are also shared with other cell types (**Fig 2C**). Yet DRD2, PTPRU, MLF2 and FMA171A1 are only identified in excitatory neurons. while RASA3, SP4, MAP3K12, ERBB4 and KCNG2 are only identified in inhibitory neurons.

Colocalization analysis for AD identifies key cell types sharing regulatory and disease risk signals (**Fig S8**). The role of microglia in genetic mechanisms of AD biology is well established, and indeed 16 genes have a colocalization signal in immune cells driven largely by microglia (**Fig 2D**). Yet, 9 genes are identified in oligodendrocytes, 12 in astrocytes and 6 in neurons at the class level, with very limited overlap between cell types. This highlights molecular mechanisms beyond microglia in AD etiology.

Analysis at the higher resolution subclass-level identifies additional genes that are not found at the class level (**Fig 2E, S7**). These genes tend to show colocalization signals in subsets of neurons since these were missed by collapsing diverse neuronal subclasses into only EN and IN classes. For example, CNTN4 coding for the contactin 4 protein involved in cell adhesion only colocalized with SZ risk in layer 6 corticothalamic excitatory neurons (EN_L6_CT) (**Fig 2F**). Similarly, SORL1 coding sortilin related receptor 1 colocalized with AD risk in microglia but not perivascular macrophages (PVM) at the subclass level or the lower resolution ‘Immune’ cell type at the class level (**Fig S8**).

### Cell-Type Specificity of Genetic Regulatory Effects Reveal Distinct Mechanisms in Neurodegenerative Disease Risk

Diverse cell types play key roles in health and disease, and characterizing differences in genetic regulatory effects at higher resolution can offer insight into the distinct functions of these cell types. While the cell type specificity of regulatory and disease biology is widely appreciated, identifying a cell type-specific regulatory effect of a genetic variant in a statistically rigorous way is challenging. Simply detecting a significant association between a genetic variant and the expression of a gene in one cell type but not another does not mean the biological effect is *specific* to the first cell type. This dilemma is common when statistical power is limited, or when there is a substantial difference in power between cell types. Indeed, widely used frequentist statistics are not adequate to address this important question ^28–30^.

We apply a multivariate Bayesian meta-analysis to produce posterior estimates of the eQTL effect size and the posterior probability that each genetic effect is non-zero ^30^. This approach shrinks effect size estimates across genes and cell types to be more robust to differences in statistical power. Examining genes with eQTLs detected in a single cell type by this Bayesian approach and intersecting with colocalization results highlights specific regulatory genetics and their role in disease biology (**Fig 3A**). For example, examining colocalization with AD shows genetic regulatory variants for BIN1 and EPHA1-AS1 are only detected in microglia, and for SERPINB1 and GALNT6 only in oligodendrocytes. The key AD gene APP, encoding the amyloid precursor protein, has a genetic regulatory signal specific to oligodendrocytes as well as another signal in astrocytes shared with other cell types. Other genes have separate eQTL signals detected in distinct cell types. INPP5D has an eQTL signal detected only in microglia and a separate signal detected only in astrocytes, while EGFR, PSD3, NALCN, TLE4 and WNT5B each have separate eQTL signals detectable in distinct cell types (**Fig 3A, S9**).

**Figure 3:**
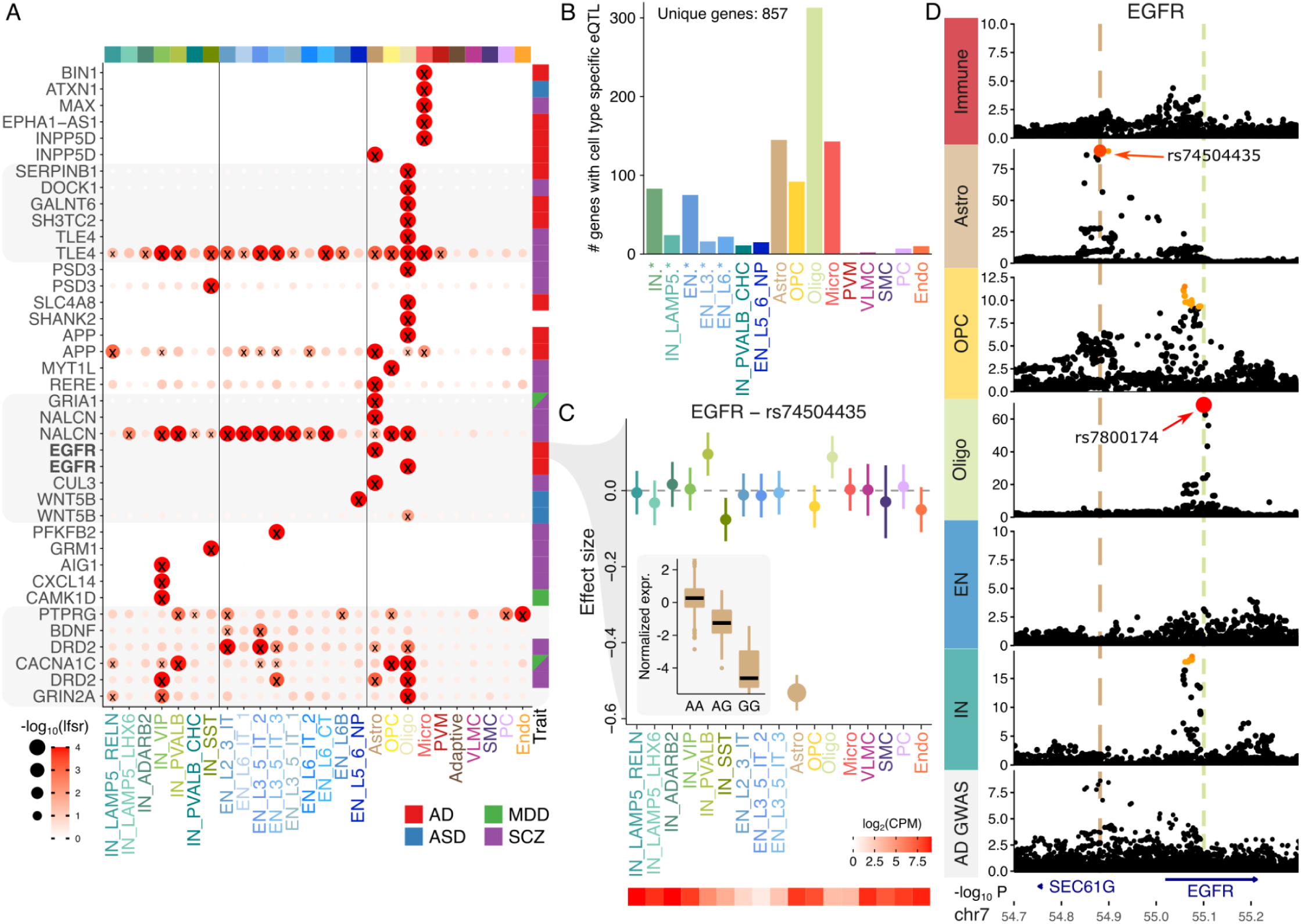
Cell type specificity of genetic regulatory effects. **A)** Cell type specific and shared genetic effects for a set of genes across each subclass. The posterior probability that the effect of the lead genetic variant is non-zero for each gene is indicated by the -log_10_ local false sign rate. Genes shown twice have independent regulatory signals in different cell types. Genes were selected based on cell type specificity, except for the bottom 6 genes were selected based on biological interest. Right column indicates genetic colocalization between eQTL and GWAS for the specified trait. **B**) Number of genes with cell type specific effects detected in each subclass or set of subclasses. Entries such as “IN.*” indicates a set of all subclasses with that prefix and the count corresponds to the number of genes with a cell type specific effect in at least one member of the set. **C**) Estimated effect size for variant rs74504435 on EGFR expression. Inset shows expression stratified by this variant in astrocytes. Error bars indicate 95% confidence interval. Heatmap (bottom) shows expression level in each subclass. **D**) Manhattan plot for EGFR showing cell type specificity for two genetic variants. Only the signal in astrocytes colocalized with genetic risk for AD. Color indicates posterior inclusion probability from statistical fine-mapping.

Using a novel composite hypothesis approach, we can directly estimate the posterior probability that a genetic regulatory effect is specific to a given cell type. Analysis at the subclass level identified 857 unique genes with cell type specific effects at posterior probability > 0.5, with 981 at the class level (**Fig 3B, S10**). At the subclass-level, oligodendrocytes have the most cell type specific genetic regulatory effects with 313 genes, followed by astrocytes with 145 and microglia with 143. While cell types with the most cell type specific findings are biologically distinct from other subclasses, neurons comprise multiple similar subclasses and indeed show less cell type specificity. We performed additional composite testing to identify genetic effects present in at least one constituent cell type.

The EGFR gene encoding the epidermal growth factor receptor has a complex signal of cell type specificity genetic regulation and colocalization with disease risk. The gene has at least two separate eQTL signals in astrocytes and oligodendrocytes. The lead variant for the astrocyte signal is rs74504435 and has a composite posterior probability of 0.946 that it is only associated with EGFR expression in astrocytes (**Fig 3C**). This genetic regulatory signal in astrocytes colocalizes with risk for AD, but the regulatory signal in oligodendrocytes is not associated with AD risk, highlighting the cell type specific role of disease biology (**Fig 3D**).

### Dynamic Genetic Regulation Across Neurodevelopment Identifies Shifting eQTL Effects and Links to Disease Risk

Neurodevelopment is a key biological process in the etiology of brain-related traits, and gene expression in some cell types changes substantially over developmental time. We hypothesized that genetic regulatory effects on gene expression also change over developmental time. Subsetting the full PsychAD dataset ^24^, Yang, et al. ^31^ extracted a neurotypical aging cohort of 284 postmortem donors age 0 to 97 comprising 1.3M nuclei and constructed a pseudotime trajectory within each cell type using a supervised method incorporating donor age ^32^. The trajectory for each cell type is anchored at early development and extends toward adulthood stages, with each nucleus assigned a continuous pseudotime value (**Fig 4A**). Dynamic eQTL effects for each cell type were detected by testing if the effect size of a genetic variant on the expression of a given gene changes along this trajectory. Analysis was performed using a negative binomial mixed model in order to consider many nuclei observed from each donor and account for overdispersion of observed counts. For example, in excitatory neurons, the genetic effect of rs1878289 on expression of NGEF, encoding a neuronal guanine nucleotide exchange factor, increases substantially during cellular maturation (**Fig 4B**). A total of 2,073 unique genes with dynamic eQTLs were detected at FDR 5%, with the number varying widely from 1,364 in excitatory neurons to only 9 in OPCs, with the highest overlap between excitatory and inhibitory neurons (**Fig 4C**). This is consistent with the extensive developmental dynamics and cellular diversity of excitatory neurons compared to the relative homogeneity of OPCs ^33^. Genes with dynamic eQTL are enriched for developmental processes like the generation of neurons and cell junction organization across multiple cell types (**Fig 4D**). Astrocytes show enrichment for nervous system processes in arterial blood pressure consistent with the key role of angiotensin production in astrocytes ^34^. Excitatory neurons show enrichment for neuronal development and differentiation, while inhibitory neurons show enrichment for neuron migration, and microglia show enrichment for axonogenesis. Oligodendrocytes are enriched for genes involved in bleb assembly, an important morphological and migratory process ^35^.

**Figure 4:**
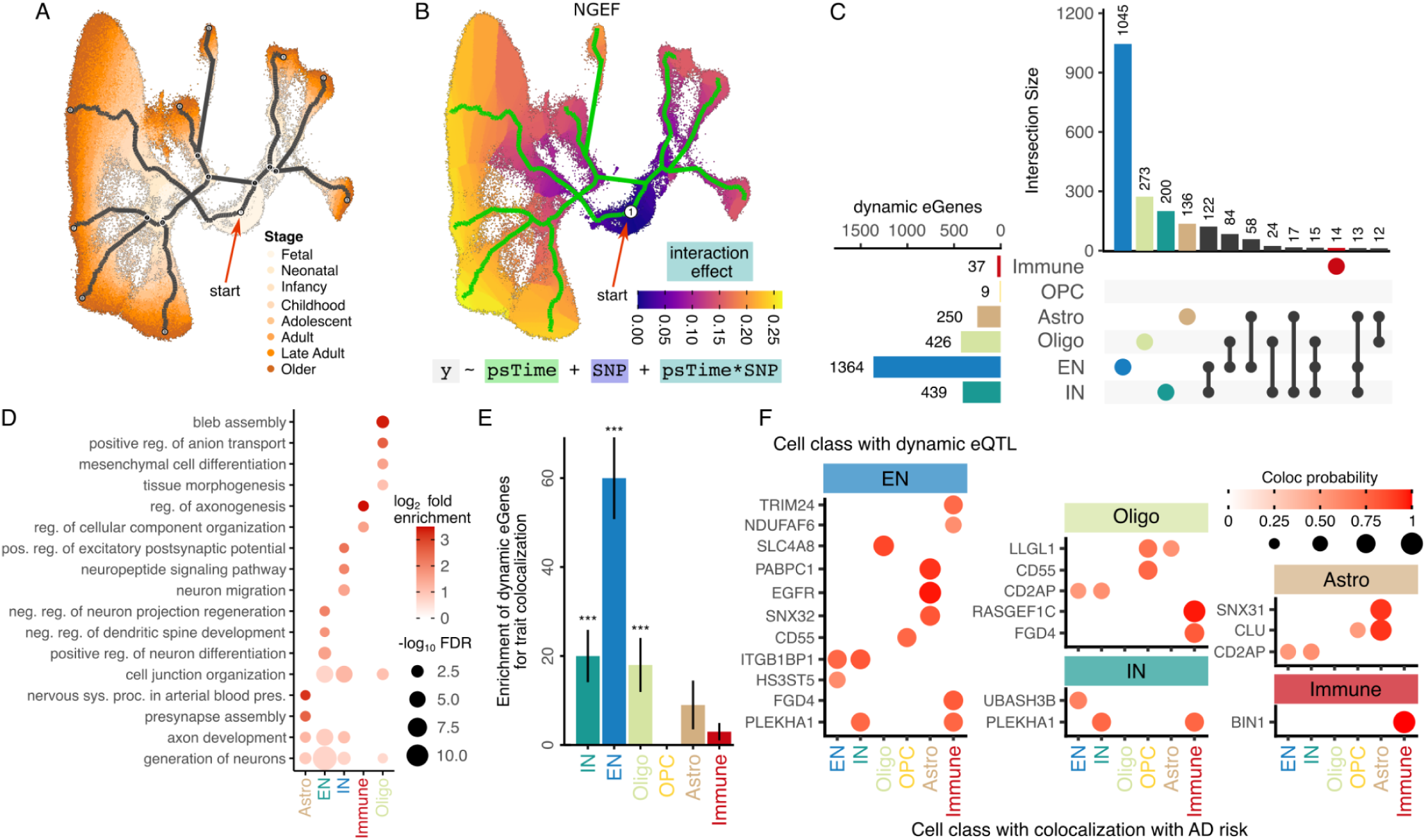
Dynamic genetic regulatory effects vary over an aging trajectory. **A)** Supervised aging trajectory in excitatory neurons with nuclei colored by developmental stage. Gray lines indicate pseudotime trajectory, red arrow indicates the beginning of pseudotime. **B**) Supervised aging trajectory in neurons from (A), with cells colored by dynamic genetic effect for NGEF changing over the pseudotime trajectory. Equation indicates regression model of gene expression (`y`), pseudotime (`psTime`), genetic variant (`SNP`) and the interaction term (`psTime*SNP`) capturing how the genetic effect changes with pseudotime. **C**) Number of dynamic eQTL detected in each cell class, along with intersection across classes. **D**) Gene set enrichment for genes with dynamic eQTLs detected in each cell class. **E**) Dynamic eGenes are enriched for genes colocalized with complex traits. **F**) Genes with a dynamic eQTL (shown in the inset of each cell class) that have a colocalization signal for AD genetic risk (shown on x-axis). For example, the gene EGFR has a dynamic eQTL detected in EN and regulatory variants for this gene colocalize with AD risk in astrocytes. Color and size of the circle indicate posterior probability from colocalization analysis.

Genes with dynamic regulatory signals detected in EN, IN and oligodendrocytes are enriched for genes with disease colocalization signals identified above, underscoring the importance of regulatory dynamics in disease biology (**Fig 4E**). Genes often have a dynamic regulatory signal detected in one cell type and a disease colocalization signal detected in a different cell type. We observe this for AD (**Fig 4F**), SCZ, MDD and ASD (**Fig S11, 12)**. This can be attributed to differences between the regulatory architecture driving dynamic changes in gene expression over aging and genetics affecting steady-state gene expression, differences in aging trajectories across cell types, as well as differences in statistical power across cell types. Meanwhile, 6 genes have a dynamic regulatory signal and disease colocalization signal detected in the same cell class (**Fig S13)**. CLU and SNX31 have a dynamic regulatory signal and colocalization signal with AD in astrocytes, ACTRB and FAM171A1 with SCZ in EN, BIN1 with AD in immune cells, and NEGR1 with MDD in oligodendrocytes.

### Trans-eQTL Mapping Identifies Brain Cell Type-Specific Genetic Regulatory Hubs and Links to Disease Risk

Genetic variants located outside the local cis-regulatory window of a gene can exert significant influence on gene expression via *trans*-regulatory mechanisms. Analysis of *trans*-regulatory signals in each cell class identifies 1655 unique genes with *trans*-eQTL signals > 5 Mb from the gene body at study-wide FDR 5%. The number of *trans*-eGenes varied by cell type, ranging from 407 in oligodendrocytes to 210 in immune cells, with limited overlap across cell types (**Fig 5A**). Analysis of genetic variants associated with multiple *trans*-eGenes identified 3 *trans*-regulatory hubs, each influencing at least three target genes, with the largest hub regulating nine downstream targets in oligodendrocytes (**Fig 5B, Fig S14**). Intersecting *trans*-eGenes with genes exhibiting cis-regulatory signals that colocalize with disease risk, we identify 12 genes with colocalization probabilities > 0.8 and 32 with colocalization probabilities > 0.5 (**Fig S15**).

**Figure 5:**
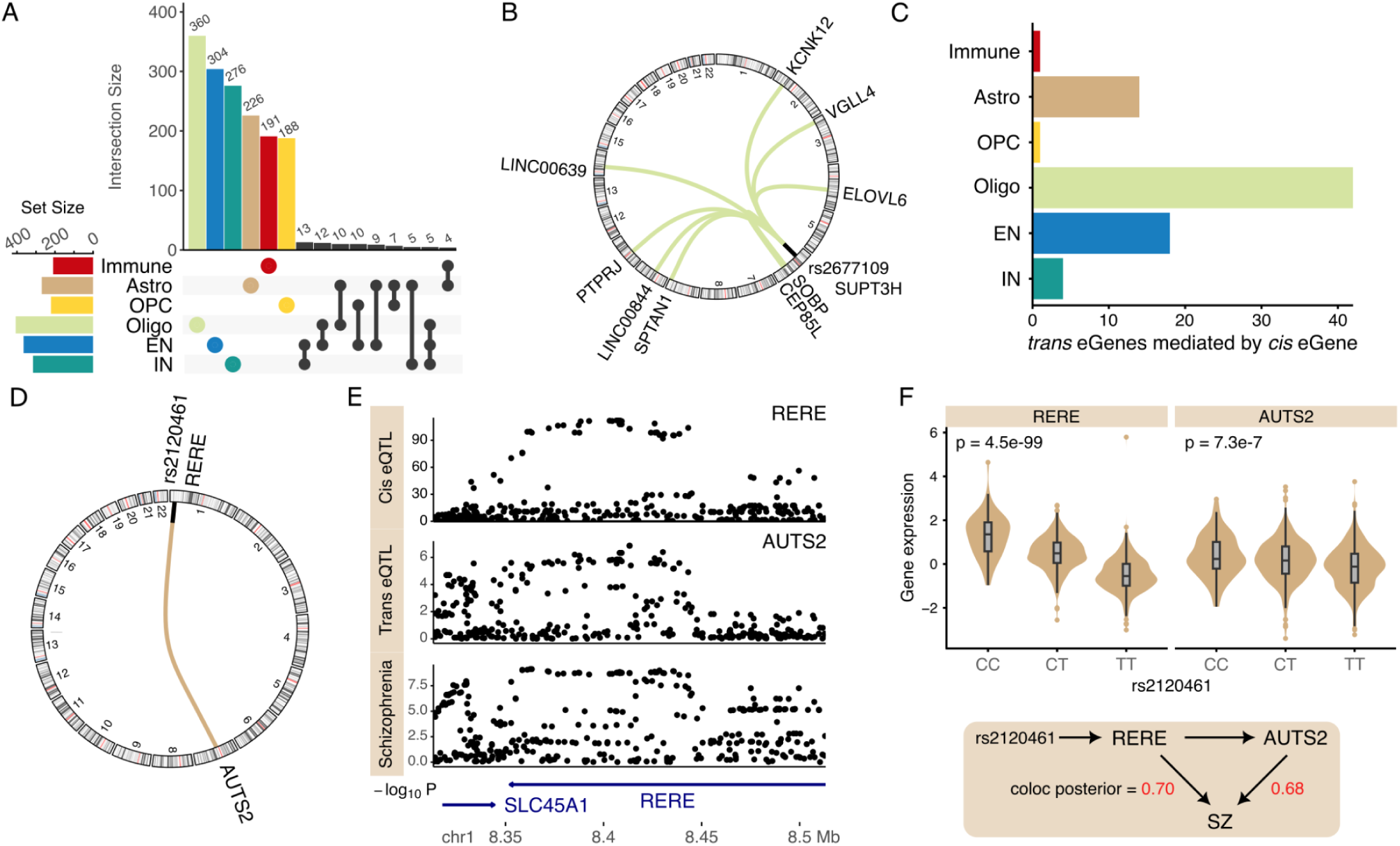
Analysis of *trans-*regulatory signals. **A)** Number of genes with *trans*-eQTLs detected in each cell class, and the overlap between classes. **B**) Circos plot of *trans*-eQTL hub showing variant rs2677109 on chr6 associated with *cis*-gene SUPT3H and with 9 *trans*-genes. **C**) Number of genes where statistical mediation analysis supports the *cis*-eQTL mediating the *trans*-regulatory signal at FDR 5%. **D**) Circos plot of *cis* association of rs2120461 and expression of RERE, and *trans* association with AUTS2. **E**) Manhattan plot showing shared genetic signal from *cis*-regulation of RERE, *trans*-regulation of AUTS2 and genetic risk for schizophrenia. **F**) Expression of RERE and AUTS2 in astrocytes stratified by rs2120461 with p-value from eQTL regression analysis (top). Proposed regulatory model where genetic regulatory variant in linkage disequilibrium with rs2120461 drives expression variation in RERE, AUTS2 in astrocytes and SZ risk (bottom). Colocalization posterior probability with SZ risk is indicated in red.

Genetic mediation analysis of these *trans-*eQTLs can identify *cis*-genes that mediate the expression of the *trans*-genes. Although analysis was underpowered to detect a large number of cis-mediators at the study-wide significance threshold of FDR 5%, using Storey’s π_1_ method ^36^, we estimate that 43% of *trans* signals are mediated by *cis*-genes, with detection primarily limited by statistical power. Nonetheless, we identified 42 *trans-*eQTLs mediated by *cis*-genes in oligodendrocytes, with fewer detected in other cell classes (**Fig 5C).**

In astrocytes, mediation analysis identified RERE as a *cis*-gene mediating the *trans*-signal between rs2120461 on chromosome 1 and expression of AUTS2 on chromosome 7 (FDR 1.5e-3) (**Fig 5D**). Both *RERE* and *AUTS2* are implicated in neurodevelopment and neuropsychiatric disorders ^37,38^. Subsequent colocalization analysis revealed that the *cis*-regulation of *RERE* and the *trans*-regulation of *AUTS2* colocalize with the genetic risk for SZ (**Fig 5E**). This suggests a mechanistic model in which the genetic regulation of RERE and its downstream effect on AUTS2 in astrocytes contribute to schizophrenia susceptibility (**Fig 5F**).

## Discussion

The brain is composed of a diverse array of cell types with distinct biology, gene expression patterns, genetic regulatory architecture, and roles in development and disease. Genetic risk variants for complex traits largely act by modifying gene expression, but our understanding of cell type-specific regulatory variation and its role in disease has been limited by sample size and cell type resolution. Here we present a multi-resolution genetic regulatory atlas of the human brain of 8 cell classes and 27 subclasses from 5.6 million single nuclei from 1384 donors of diverse ancestry. We identify cis-eQTLs for 14,258 genes, and observe wide variation in the number of cis-QTLs detected for each cell class and subclass due to differences in cell type abundance. This highlights the importance of increasing sample size in order to study the regulatory architecture of rarer cell types. The regulatory signals are distinct across cell types and, for non-neuronal cells, reflect chromatin accessibility in each cell type.

Integration with genome-wide association studies for brain-related traits identifies cell types mediating genetic risk for each trait. Colocalization analysis further identifies specific genes and cell types mediating genetic risk that add novel insight into disease biology. While the role of neurons in schizophrenia (SCZ) is well established ^39^, the role of neuron subtypes is not well understood. As a step towards a higher resolution understanding, we identified multiple genes colocalizing with SZ risk in specific neuronal subtypes. While the role of genetic regulation of the CNTN4 expression in SZ was first identified from bulk gene expression profiling ^12^, we find a shared regulatory and SZ risk signal only in layer 6 corticothalamic excitatory neurons.

Recent work in Alzheimer’s disease (AD) has uncovered the unique role of microglia, brain-resident myeloid cells, in genetic risk and molecular etiology ^40,41^. In addition to identifying genetic regulatory signals shared with AD risk in microglia, we also identify genes that are not detected in microglia. Among others, these include well-studied genes like APP, SNX31, SNX32, EGFR and CLU in astrocytes; CR1 and CR2 in oligodendrocytes; CTSB, CTSH, and ACE in neurons.

Identifying cell type-specific regulatory effects, where a genetic effect is non-zero in only a specified cell type, is challenging with existing frequentist methods. Building on Bayesian multivariate meta-analysis, we prioritize genes based on their specific- versus shared-regulatory signals and examine the complex regulatory architecture that is active in specific cell types. We highlight the example of EGFR, which has at least two distinct regulatory programs, with one active in astrocytes and another in oligodendrocytes. Notably, only the regulatory signal in oligodendrocytes colocalizes with the genetic risk of AD, highlighting the importance of cell type-specific regulatory programs in disease biology.

Early developmental processes play a key role in neurodevelopmental and neuropsychiatric disease ^42^. Yet studying the genetic regulatory architecture at cell-type resolution at this key stage is especially challenging. Here we leverage the wide age span of donors in this dataset and integrate with a neurotypical aging cohort from the PsychAD to construct a pseudotime trajectory within each cell class. We identify dynamic eQTLs by testing for genetic effects that vary over developmental time. Dynamic genetic effects are most prevalent in excitatory and inhibitory neurons, and dynamic eGenes in these classes are enriched for colocalization with brain-related traits.

The unique scale of this dataset enabled the discovery of trans-eQTLs for the most abundant cell classes. We find limited overlap between cell classes, and we identify trans-eGenes that also have a *cis*-regulatory signal colocalizing with disease risk. In astrocytes we identify a cis-eQTL for RERE as a trans-eQTL for AUTS2 and both regulatory signals colocalized with genetic risk for SZ, underscoring the role of complex trans-regulatory architecture in disease biology.

Our findings provide key insights into the cell type-specific genetic regulation underlying neuropsychiatric and neurodegenerative diseases. This study underscores the importance of expanding sample sizes and increasing single-cell resolution to capture rarer cell types, paving the way for a deeper understanding of disease mechanisms. As we move forward, integrating multi-omic data and ensuring representation of diverse ancestries will be critical to advancing precision medicine and developing targeted therapeutic strategies for brain disorders.

## Methods

### Sample selection and preprocessing

Brain tissue from the dorsolateral prefrontal cortex was obtained from 1,494 donors by the PsychAD Consortium ^24,25^. The dataset comprised donors from 3 sources. The Rush Alzheimer’s Disease Center (RADC) repository of tissue from the Religious Orders Study or Rush Memory and Aging Project ^43^ provided 152 specimens, Human Brain Collection Core (HBCC) provided 300, and Mount Sinai Brain Bank (MSSM) provided 1,042 contributors. The cohort includes similar numbers of males and females, and spans the entire postnatal age range of 0 to 108. For additional details about the donors and data processing see Lee, et al. ^24^ and Fullard, et al. ^25^.

Paired-end reads from snRNA-seq libraries were aligned to the hg38 reference genome using STAR solo ^44^, and sample pools were demultiplexed through genotype matching with vireo ^45^. Following the generation of per-library count matrices, downstream processing was conducted using Pegasus v1.7.0 ^46^ and scanpy v1.9.1^47^. We implemented a stringent QC process to eliminate ambient RNA and preserve high-quality nuclei for further analysis.

Genomic DNA was extracted from frozen brain tissue using the QIAamp DNA Mini Kit (Qiagen) following the manufacturer’s instructions. The samples were genotyped using the Infinium Psych Chip Array (Illumina) at the Mount Sinai Sequencing Core. Pre-imputation processing involved running the quality control script HRC-1000G-check-bim.pl from the McCarthy Lab Group, utilizing the Trans-Omics for Precision Medicine (TOPMed). Genotypes were phased and imputed on the TOPMed Imputation Server (https://imputation.biodatacatalyst.nhlbi.nih.gov). Samples were excluded if there was a mismatch between self-reported and genetically inferred sex, suspected sex chromosome aneuploidies, high relatedness (KING ^48^ kinship coefficient > 0.177), or outlier heterozygosity (± 3 SD from the mean). Additionally, samples with a sample-level missingness > 0.05 were excluded, as calculated within a subset of high-quality variants (variant-level missingness ≤ 0.02). A subset of 1,384 donors with genotype data were analyzed in this study.

### Normalization of gene expression

Pseudo-bulk read counts were calculated by summing reads from the same individual using the dreamlet workflow ^49^. We regressed out the effect of the sample pool and the proportion of mitochondrial expression from each cell to remove batch effects, and the influence from disease status was also removed by regression. Finally, the residuals were divided by the predicted standard deviation to produce Pearson residuals, excluding the impact of varying sequencing depths among the scRNA-seq libraries.

We applied PEER package ^50^ to detect hidden unobserved covariates. To find an optimal number of PEER factors to remove, we performed eQTL detection called on the input expression matrix normalized by pre-selected biological and technical covariates but differing in the numbers of PEER factors between 10 and 98 with an interval of 4. The setting with the most genome-wide significant eQTL detected was used as the final results for downstream analysis.

### Analysis of genetic regulatory variants at pseudobulk level

In each cell class and subclass, eQTL analysis was performed for variants within 1 Mb of the transcription start site of each expressed gene. The mmQTL software was used to fit a linear mixed model to account for the diverse genetic ancestry of the donors in this dataset ^7^.

### Evaluating replication of genetic regulatory variants across datasets

We used the R package qvalue to estimate eQTL replication rates using Storey’s π_1_ statistic ^36^. For a pair of datasets, we first extracted the most significant variant for genes with eQTL in the discovery data. The p-values from the replication dataset were then used to estimate Storey’s π_1_ value, which indicates the fraction of hypothesis tests for which the null is rejected. Thus, π_1_ is the estimated fraction of eQTLs that replicate in the second dataset. This metric of replication is useful because it does not depend on hard cutoffs for p-values for FDR, and it has been widely adopted.

### Enrichment of open chromatin regions around detected regulatory variants

To determine if cell type-specific regulatory elements were enriched around eQTLs, we used the fdensity function in QTLtools ^51^ to compute the number of functional elements that overlap each 10-kb bin in a 2 Mb window around the *cis*-eQTL. The open chromatin region (OCR) annotations were obtained from single cell ATAC-seq from human brain ^26^, and cell type specific OCRs were defined as those found in only one cell type.

### Partitioning heritability based on statistical fine-mapping

Stratified LD score regression (S-LDSR) was used to test if custom variant annotations from statistical fine-mapping of eQTL signals were enriched for heritability for genetic risk for complex traits ^3,52^. For each eGene, statistical fine-mapping was used to compute a posterior inclusion probability for each *cis*-variant and retaining variants in the 95% credible set for each gene. Each variant in the genome is annotated with a probability value from this analysis. Variants not in a 95% credible set receive a value of 0, and variants evaluated for multiple genes receive the maximum probability value for the variant across these genes. This approach is termed ‘MaxCPP’ in Hormozdiari, et al. ^52^. S-LDSC was then used to partition trait heritability using the constructed functional annotations. The estimated enrichment was used to measure the importance of each eQTL category to human complex traits or diseases. To rule out the potential influences of the correlation among eQTL categories, we aggregated the baselineLD model, which includes a set of 75 functional annotations, to create functional annotations for the eQTL category and ran S-LDSR jointly.

### Proportion of disease heritability mediated by regulatory variants

Mediated expression score regression (MESC) estimates the proportion of disease heritability mediated by regulatory variants for a specific set of molecular traits ^27^. We applied this approach to estimate the contribution of regulatory variants across cell classes and subclasses to the heritability of complex traits. MESC was then used to calculate mediated heritability with default settings. To estimate the joint contribution of subtypes from one cell class, we also run meta-analyzed MESC by running meta_analyze_weights.py

### Colocalization of genetic signals from regulatory and disease risk variants

To evaluate the relationship between molecular QTL, we used the coloc R package to conduct colocalization analysis ^53^. The summary results from meta-analysis were used as input for coloc. The phenotypic variance was set to 1 as we have normalized the summary results before meta-analysis, but otherwise, parameters were set to be default values. We also applied an extension, moloc ^54^, that applies this framework to identify the colocalization of 3 signals. For coloc analyses, we considered signals between two traits to be colocalized at posterior probability ≥ 0.5 (i.e pp4 ≥ 0.5).

### Identifying shared and cell type specific genetic regulatory effects

To determine how eQTL effects are shared between different cell types, we applied a multivariate Bayesian meta-analysis approach using the mashr software ^30^. The software uses a Bayesian approach to shrink effect sizes across genes and cell types in order to estimate the posterior effect sizes and posterior probability that an effect has the correct sign. Following guidelines in the mashr documentation, we estimated the prior effects size distribution using a collection of genes expressed in all cell types and learned the empirical correlation structure using 600k randomly selected variant-trait pairings. For all genes with a genome-wide significant eQTL in at least 1 cell type, the variant with the smallest p-value was selected and the coefficient estimate and standard error were used in analysis with mashr. For genes not analyzed in a particular cell type due to insufficient expression, values of 0 were used for the coefficient and 1e6 were used for the standard error.

We extend this approach to develop a formal statistical test to identify cell type specific genetic regulatory effects. Although mashr analysis integrates results across cell types, the software characterizes the regulatory effect of a variant in one cell type at a time. Mashr analysis asks if there is genetic effect in a given cell type. Instead, we directly test if a genetic effect is cell type specific using a composite test to ask if a genetic effect is non-zero in a given cell type while also being zero in all other cell types.

Here we describe the math of the composite test. For a gene j and cell type i, mashr reports the local false sign rate defined as lfsr*_j,i_* = *min* [*p*(*β_i,j_* ≥ 0|*β̂*, …)*p*(*β_i,j_* ≤0|*β̂*,…)], where is *p*(*β_i,j_* ≤0|*β̂*,…) the probability that the true value of the regression coefficient is greater than zero given the estimated coefficient and other model parameters ^55^. Thus lfsr*_j,i_* is the posterior probability that the sign of the estimated coefficient does not agree with the sign of the true coefficient value, and is more conservative than the local false discovery rate^55^. Then let *p_i,j_* =1 – lfsr*_j,i_* be the probability that the signs agree. For a given gene, this set of posterior probabilities can be used to estimate the probability of any combination of eQTL effects across cell types. Therefore, *p_i,j_* is taken as a conservative estimate of the probability that a genetic variant has a non-zero effect size in cell type 1, and 1 – *p*_2,*j*_ is taken as a conservative estimate that the effect is zero in cell type 2. Combining these, the probability of a non-zero effect in cell type 1 and a zero effect in cell type 2 is *p*_1,*j*_(1 – *p*_2,*j*_), assuming the probabilities are independent. In general, the probability of an arrangement with non-zero effects in set 1 all zero effects in set 2 is [Π*_i_*_∈set1_ *p_i,j_*][Π*_i_*_∈set2_(1 – *p_i_*_,*j*_)].

Due to limited statistical power to detect eQTLs in high resolution cell types, it is often too restrictive to ask, for example, if an eQTL effect is non-zero in all excitatory neuron subtypes. Instead, we can ask if there is a non-zero effect in at least one subtype by evaluating1 - [Π*_i_*_∈set1_(1 – *p_i_*_,*j*_)]..

### Aging-related dynamic-eQTL detection

A subset of the full PsychAD dataset ^24^ was extracted by Yang, et al. ^31^ to create a neurotypical aging cohort of 284 postmortem donors age 0 to 97 comprising 1.3M nuclei. Yang, et al. ^31^ then constructed a pseudotime trajectory within each cell type using a supervised method incorporating donor age by applying the UMAP of MATuration (UMAT) method that restricts UMAP neighbor selection to nuclei from nearby developmental stages ^32^. Each nucleus is assigned a pseudotime score, and we used a regression approach to statistically test if the regulatory effect of a variant in a given gene expression trait changes along the developmental trajectory. The gene expression counts for each nucleus were the response and the pseudotime, genetic variant and an interaction between the two were predictors in the model. Covariates for library size, age, sex and mitochondrial rate were included as fixed effects. Since many nuclei are observed for each donor, the donor was included as a random effect. We used a negative binomial mixed model to model overdispersion of the count data to protect against false positives. Analyses were implemented using the glmer.nb() function in the lme4 R package ^56^. P-values were extracted from a Wald test.

The enrichment of genes with dynamic regulatory signals for colocalization with disease traits was evaluated as follows. For a cell class with dynamic eGenes, the number of genes that also have a colocalization signal was computed. The null distribution of this count was evaluated by randomly sampling genes and evaluating the overlap with genes with a colocalization signal. The standard error of the overlap for each cell type was evaluated using 100 rounds of random sampling.

### Trans-eQTL detection

We used 56,204 eSNPs from the *cis*-eQTL analysis and 45,088 genome-wide significant variants from 8 brain disease GWAS studies to map trans-eQTLs, and the normalized gene expression from cell-class level were used as phenotype. SNP genotype was included as the dependent variable for all gene-variant pairings in a linear regression model that we tested. We defined trans-variants as variants that are beyond 5 Mb distance to target genes, and focused on autosomal chromosomes, and excluded any signal within MHC regions. Genes with a mappability score < 0.8 were excluded to avoid false positive trans-eQTL findings due to reads mapping to multiple locations in the genome ^10,57^. Multiple testing correction was first applied to each gene separately by using a Sidak corrected p-value according to where is the vector of p-values for each variant tested for a given gene, and is the number of tests. We used . A second round of multiple testing correction was applied across all genes and cell types by using the Benjamini-Hochberg method ^58^ on these Sidak-corrected p-values. Study-wide significant trans-eQTL were identified at FDR 5%.

### Mediation analysis to explore *cis*-mediation *trans*-eQTL

In order to find trans-eQTL demonstrating evidence of mediation, we limited our exploration to trans-eSNPs with high LD (r^2^ ≥ 0.75) with at least one peak eQTL variant within 1Mb. We applied the strategy developed in Pierce et al, ^59^ to calculate the indirect impact of trans-SNP on trans-eGenes. Multiple test correction based on the BH approach was applied to control the study-wide FDR at 5%.

## Data Availability

All data produced in the present study are available upon reasonable request to the authors

## Code availability

mmQTL, https://github.com/jxzb1988/mmQTL

dreamlet: https://diseaseneurogenomics.github.io/dreamlet/

All the source codes utilized in this study are available at https://github.com/DiseaseNeuroGenomics/nps_ad

## Acknowledgments

We would like to express our deep gratitude to the patients and their families who generously donated the invaluable biological material essential for the success of this study. We are profoundly indebted to their participation and commitment to advancing scientific knowledge and improving human health. We acknowledge the National Institute on Aging for their generous support in funding this research with the following NIH grants: R01AG067025 (PR), R01AG082185 (PR), R01AG078657 (GV), R01AG065582 (PR), R01MH125246 (PR). Human tissues were obtained from the NIH NeuroBioBank at the Mount Sinai Brain Bank (MSSM; supported by NIMH-75N95019C00049), the Rush Alzheimer’s Disease Center (RADC; funding: P30AG10161, P30AG72975, R01AG15819, R01AG17917, R01AG22018, U01AG46152, and U01AG61356), and NIMH-IRP Human Brain Collection Core (HBCC, project # ZIC MH002903). The results published here are in whole or in part based on data obtained from the AD Knowledge Portal. This work was supported in part through the computational and data resources and staff expertise provided by Scientific Computing and Data at the Icahn School of Medicine at Mount Sinai and supported by the Clinical and Translational Science Awards (CTSA) grant UL1TR004419 from the National Center for Advancing Translational Sciences. Research reported in this publication was also supported by the Office of Research Infrastructure of the National Institutes of Health under award number S10OD026880 and S10OD030463. The content is solely the responsibility of the authors and does not necessarily represent the official views of the National Institutes of Health.

## Author contributions

Conceptualization and study design: B.Z., G.E.H., P.R.. Data contribution or analysis tools: H.Y., P.F.N.U., D.M., P.A., D.A.B., S.M., V.H., G.V., D.L., J.F.F., J.B., K.G., G.E.H., P.R. B.Z., H.Y., G.E.H. performed the analyses. B.Z., J.F.F., G.E.H., P.R. wrote the manuscript with input from all authors.

