## Supplementary material for "Single-Nucleus Atlas of Cell-Type Specific Genetic Regulation in the Human Brain": FigureS1 to FigureS15

### Supplementary Figures

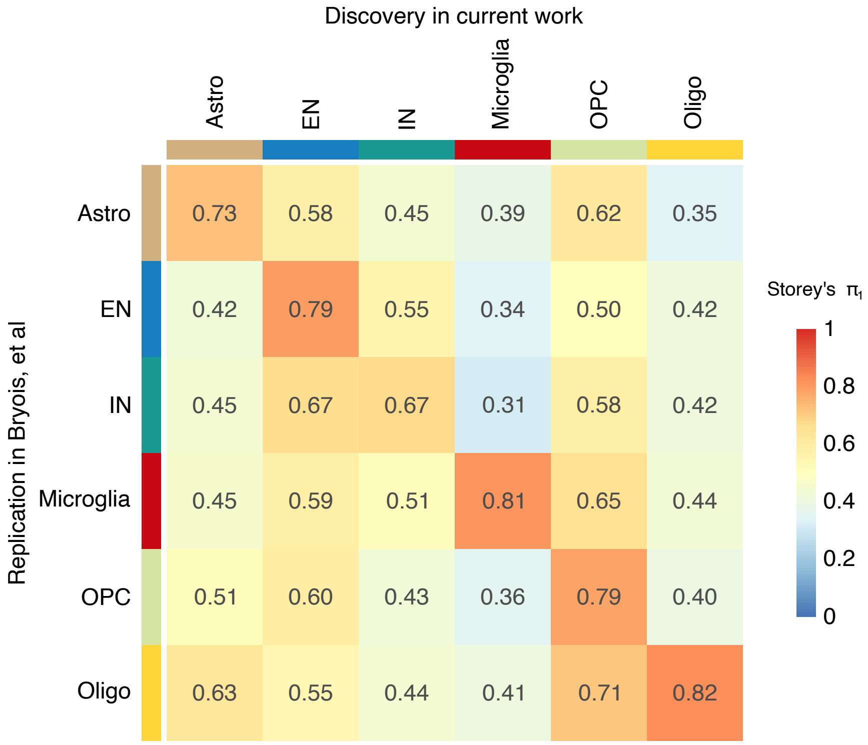


**Figure S1: Replication rate of regulatory variants from each cell class estimated by Storey's $\pi_1$.** The current dataset (columns) is used for eQTL discovery and the replication rate is evaluated in Bryois, et al. (2022) (rows). For example, lead eQTL variants discovered in astrocytes in the current work were replicated in astrocytes in at a rate if 73%, but only 42% in EN. The fact that the strongest replication rate is seen along the diagonal for the matching cell type indicates replication of the specific regulatory architecture of each cell type.

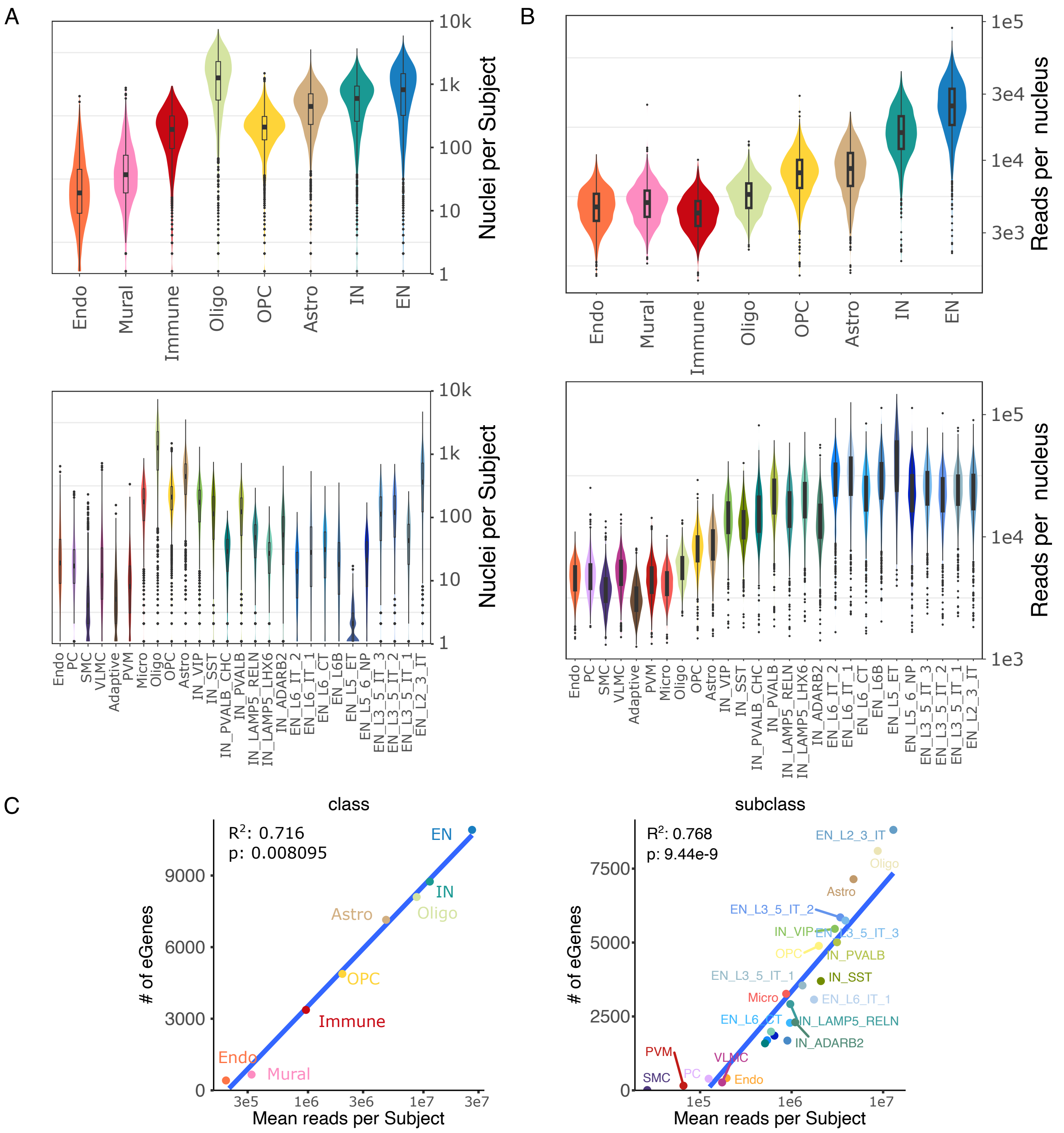


**Figure S2: Relationship between number of cells, reads and eQTL detection A)** Number of nuclei per subject for class (top) and subclass (bottom). **B)** Number of reads per nucleus for class (top) and subclass (bottom). **C)** The number of genes with significant eQTLs (i.e. eGenes) detected in each class (left) or subclass (right) shown as a function of the mean reads per subject for each cell type. Blue line indicates least squares fit. Squared Pearson correlation and p-value from linear regression are shown.

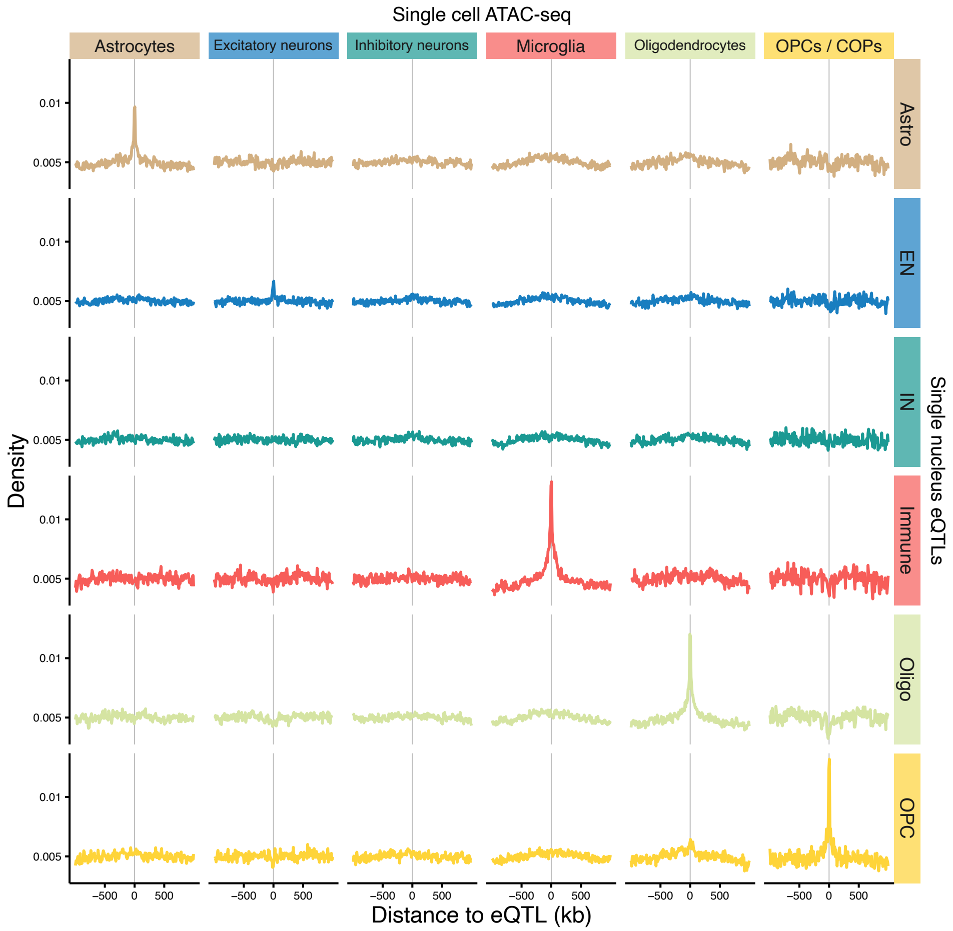


**Figure S3: Enrichment of lead eQTL variants near open chromatin regions.** Enrichment is shown for eQTLs detected in each cell class (rows) for open chromatin regions detected in each cell type from single-cell ATAC-seq (columns). Cell type annotations from Corces et al. (2020) are used for ATAC-seq data. 'COP' indicates committed oligodendrocyte precursor cells

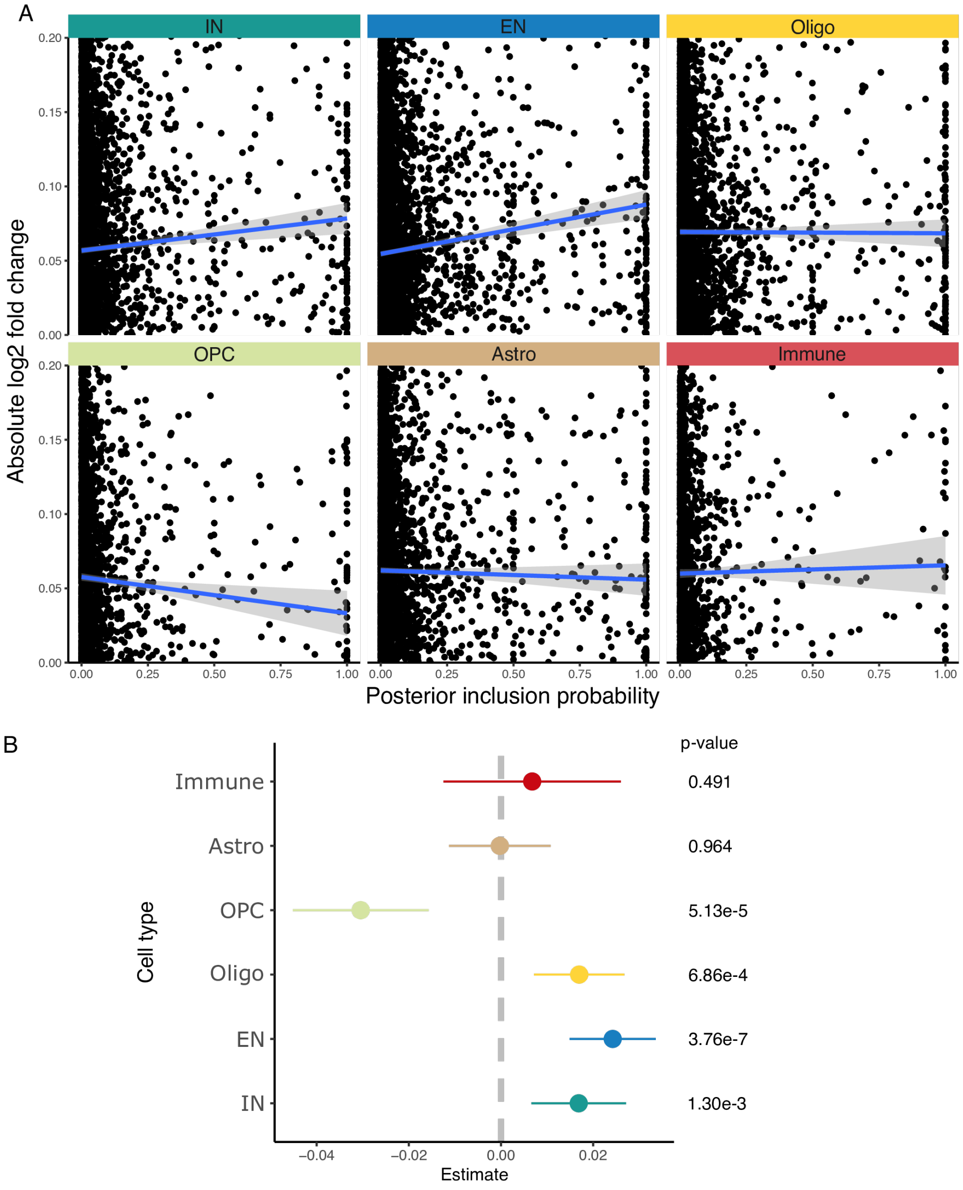


**Figure S4: Integration with allelic affects from multiplexed parallel reporter assay (MPRA). A)** Relationship between allelic effects from MPRA and fine-mapping posterior probability from each cell class. The blue line indicates fit with linear model, and the gray band shows standard error. Although some SNPs are excluded from this plot because of the y-axis range, all SNPs were used to fit the regression model. **B)** Regression coefficient for the slope from (A) indicates allelic effect size from MPRA is most associated with fine-mapping posterior probability in excitatory neurons (EN). Errors bars indicate 95% confidence interval. P-value for each hypothesis test is indicated on the right.

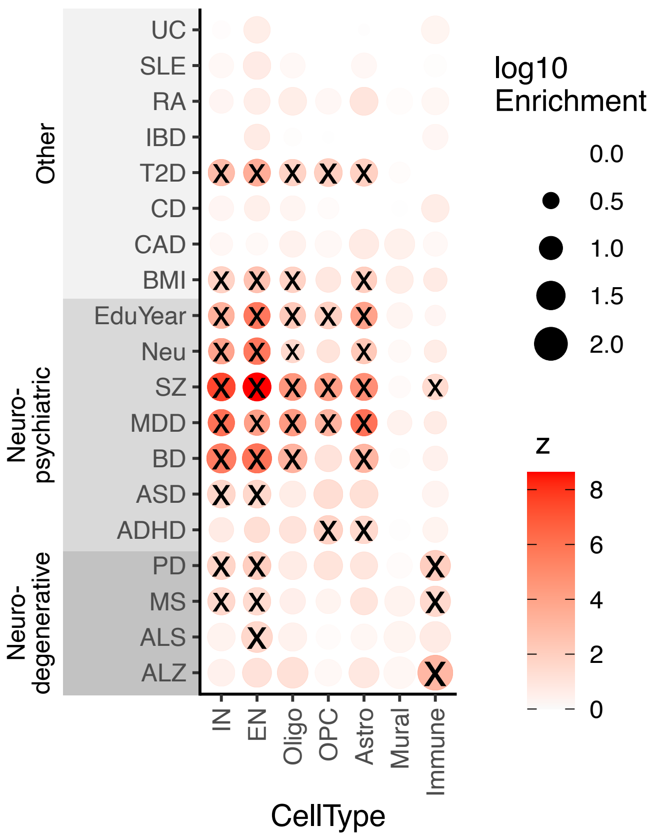


**Figure S 5: Regulatory variants are enriched for heritability of complex traits.** Enrichment of genetic regulatory variants in the 95% credible set from statistical fine-mapping for heritability of genetic traits. Color indicates z-statistic of null hypothesis of no enrichment, point size indicates log10 enrichment. Tests with FDR $<$ 5% are indicated by ‘X’.

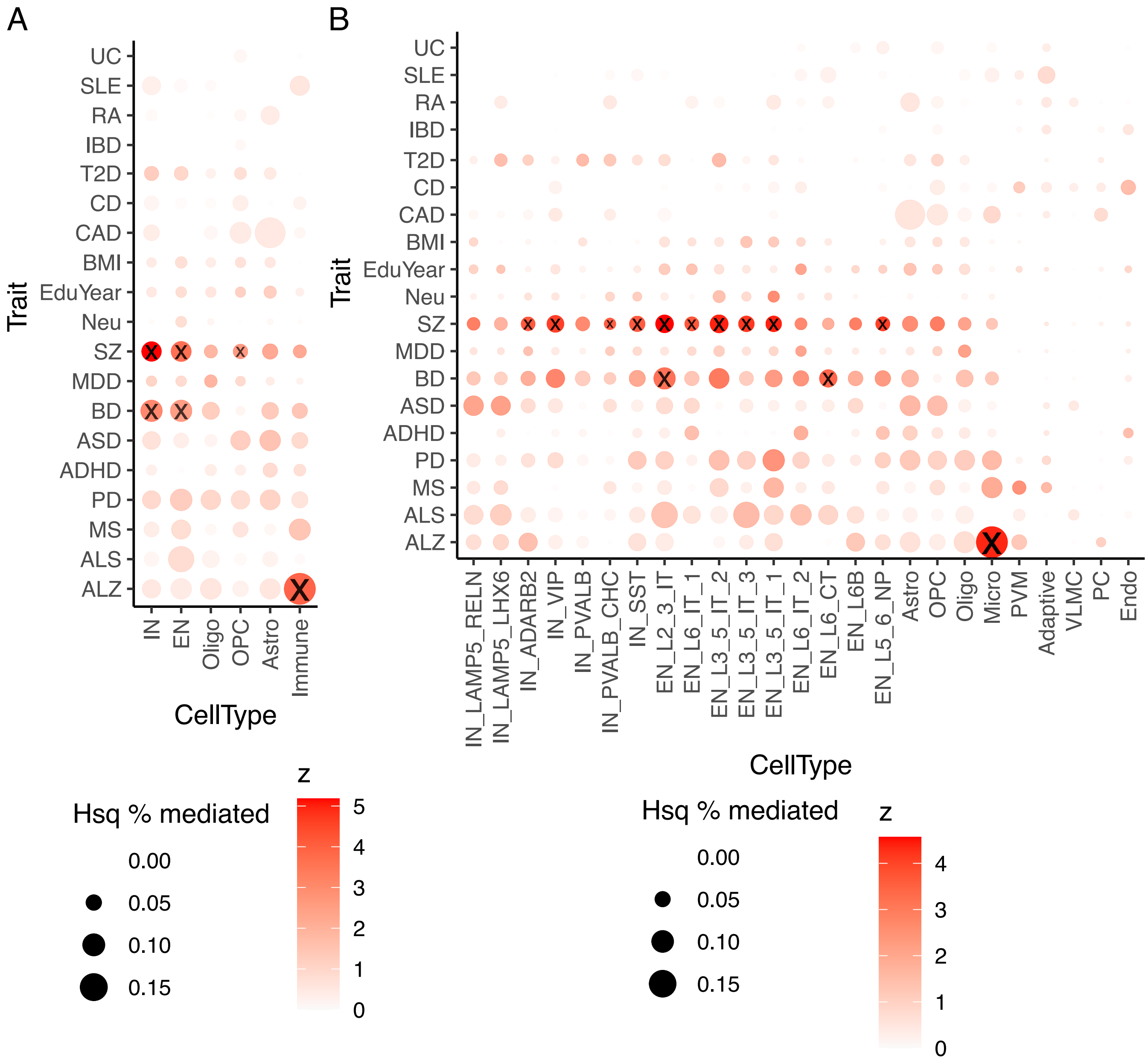


**Figure S 6: Mediation of trait heritability**. Fraction of heritability of complex traits mediated by regulatory variants at the **A)** class and **B)** subclass levels. Tests with FDR < 5% are indicated by 'X'.

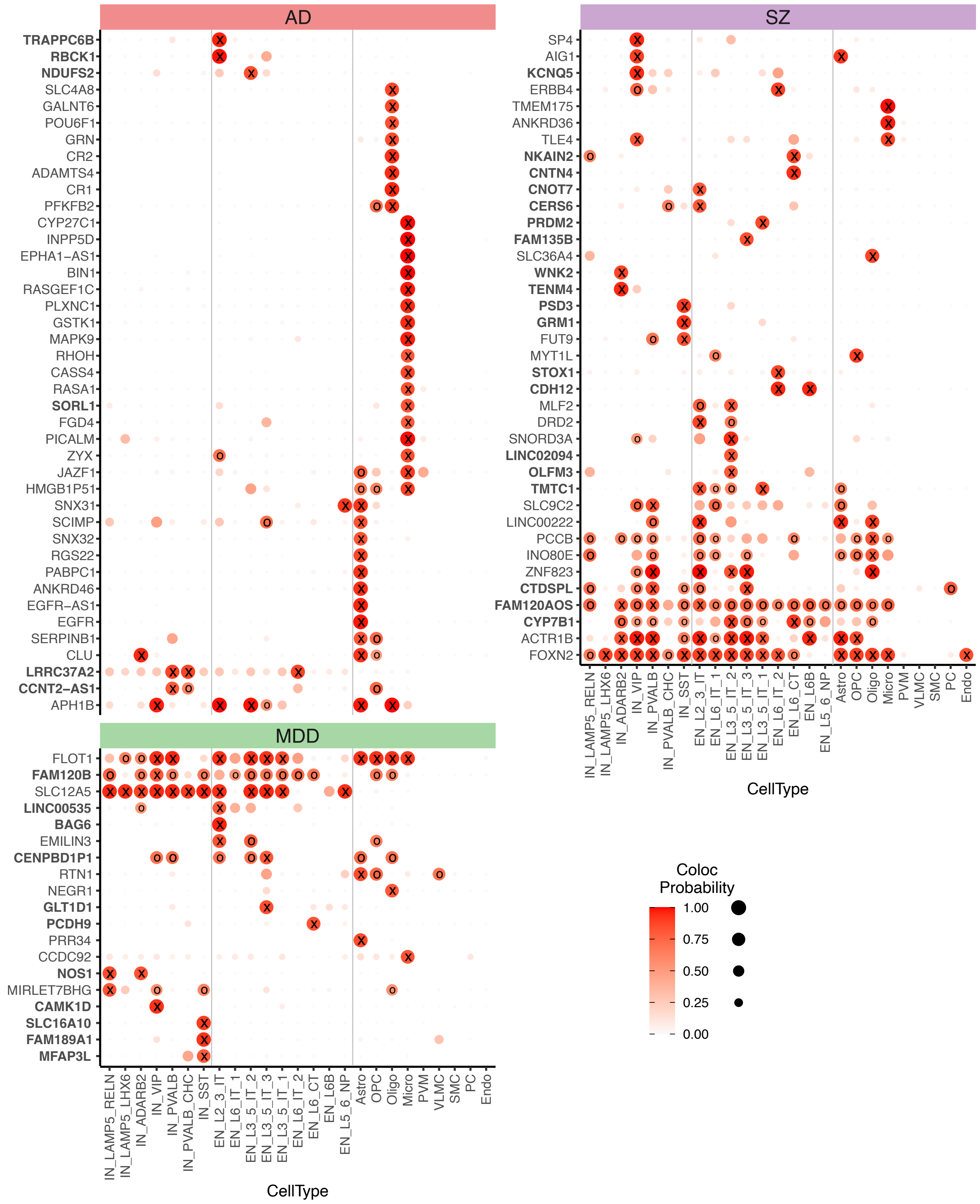


**Figure S7: Colocalization signal at the subclass-level**. Posterior probabilities $> 0.8$ are indicated by 'X', and $> 0.5$ are indicated by 'o'. Genes in bold do not have colocalization posterior probability $> 0.8$ at the class level.

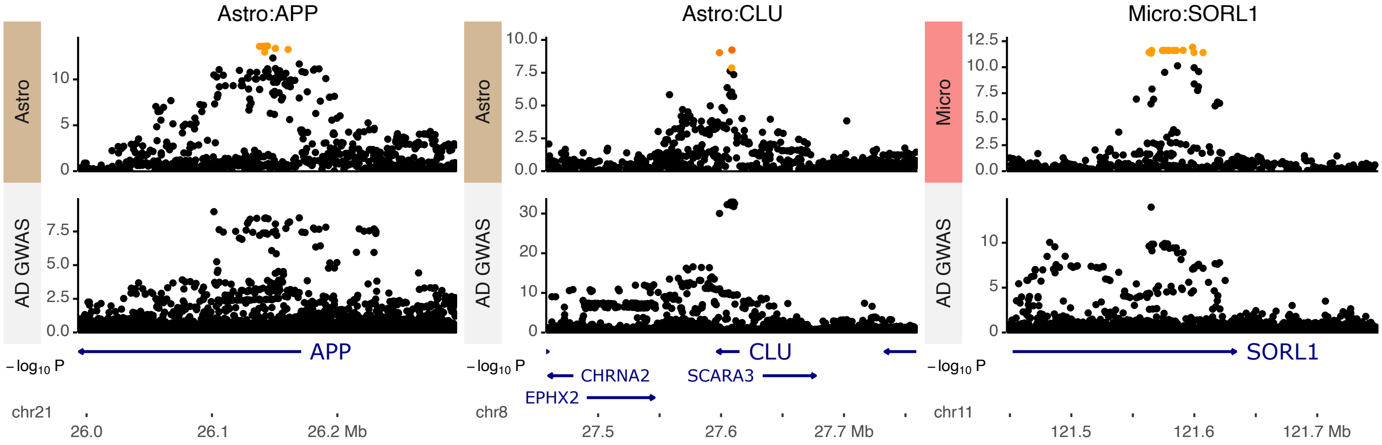


**Figure S8: Manhattan plots for colocalization signals.** Colocalization with AD genetic risk is observed for APP in astrocytes (left), CLU in astrocytes (center), and SORL1 in microglia (right).

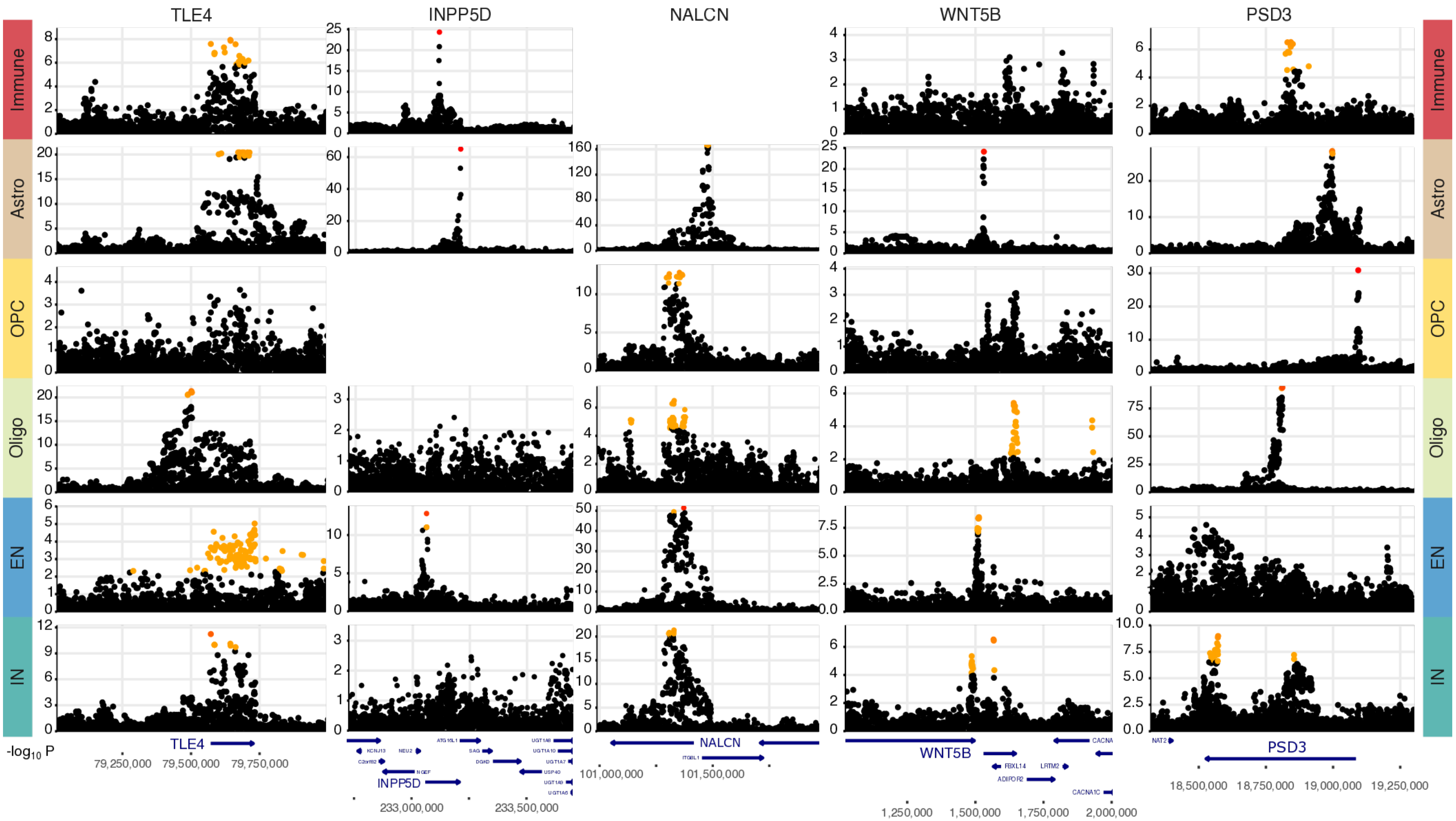


**Figure S9: Manhattan plot of genes with independent eQTL signals in different cell types.** Color indicates posterior inclusion probability from statistical fine-mapping. Blank panels indicates case where the gene is not sufficiently expressed in the given cell type.

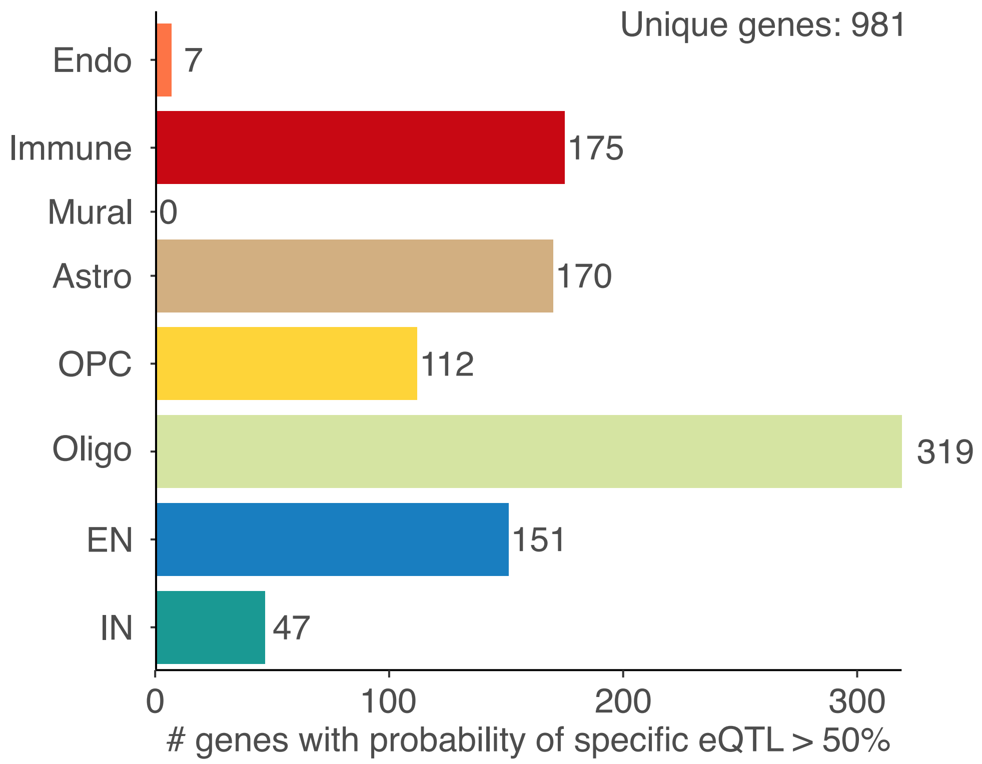


**Figure S10: Cell type specific findings at the class-level using multivariate Bayesian meta-analysis followed by composite test.**

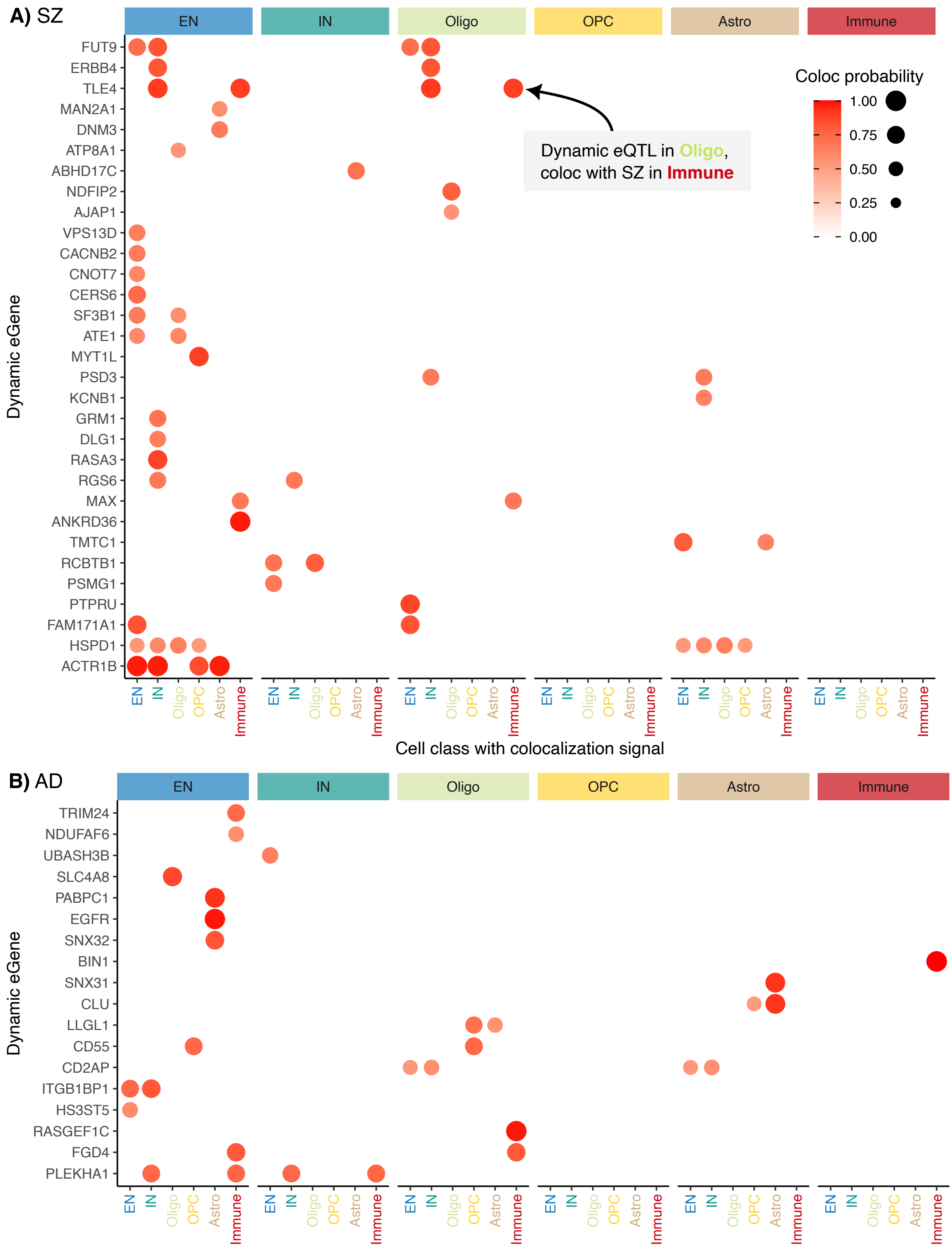


**Figure S11: Genes with a dynamic eQTL and a disease colocalization signal.** Genes with a dynamic eQTL (shown in inset of each cell class) that have a disease colocalization signal (shown on x-axis). For example, the gene TLE4 has a dynamic eQTL detected in oligodendrocytes and regulatory variants for this gene colocalize with AD risk in immune cells. Color and size of circle indicates posterior probability from colocalization analysis. Results are shown for **A**) SZ and **B**) AD.

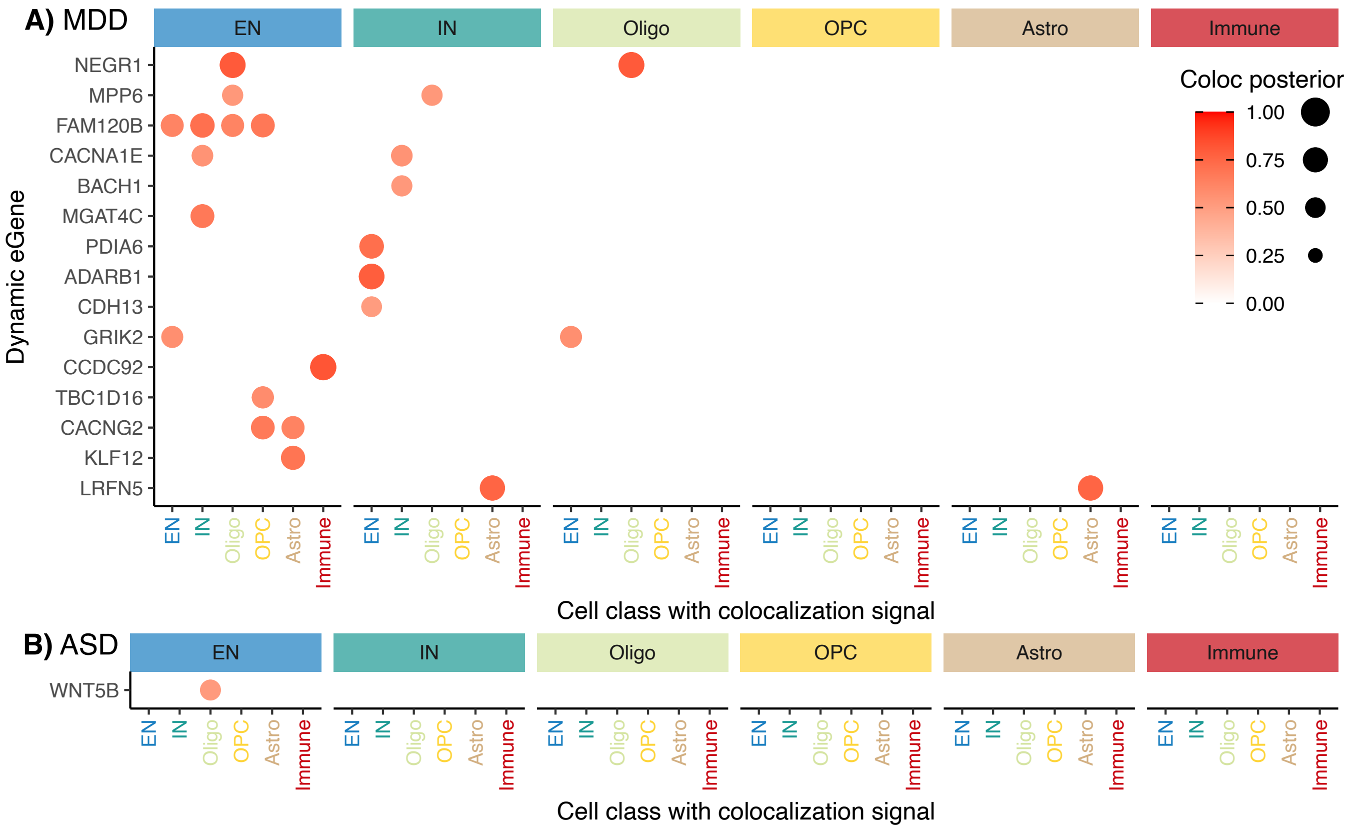


**Figure S12: Genes with a dynamic eQTL and a disease colocalization signal.** Genes with a dynamic eQTL (shown in inset of each cell class) that have a disease colocalization signal (shown on x-axis). For example, the gene TLE4 has a dynamic eQTL detected in oligodendrocytes and regulatory variants for this gene colocalize with AD risk in immune cells. Color and size of circle indicates posterior probability from colocalization analysis. Results are shown for **A**) MDD and **B**) ASD.

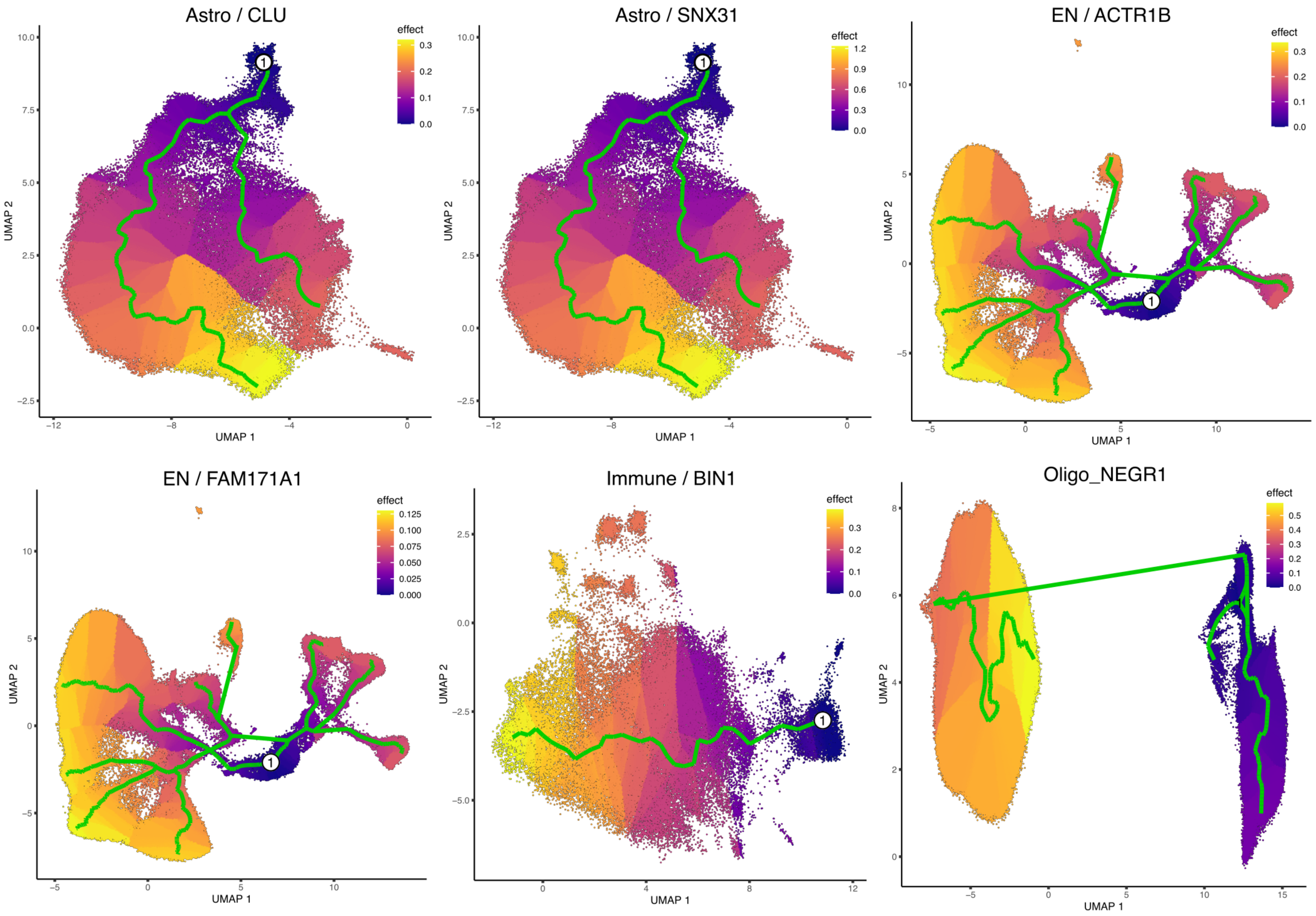


**Figure S13: Dynamic eQTLs at the class-level.** Supervised aging trajectories with cells colored by dynamic genetic effect changing over the pseudotime trajectory.

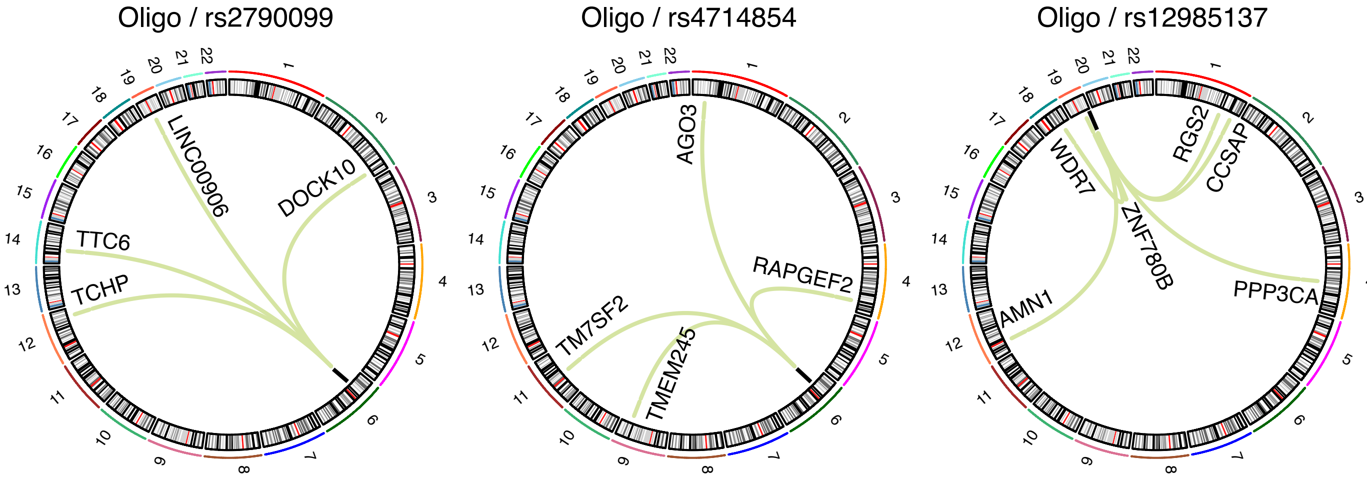


**Figure S14: Trans-regulatory hubs.** Trans-regulatory hubs with more than 2 downstream target genes at a study-wide FDR of 5% were identified only in oligodendrocytes.

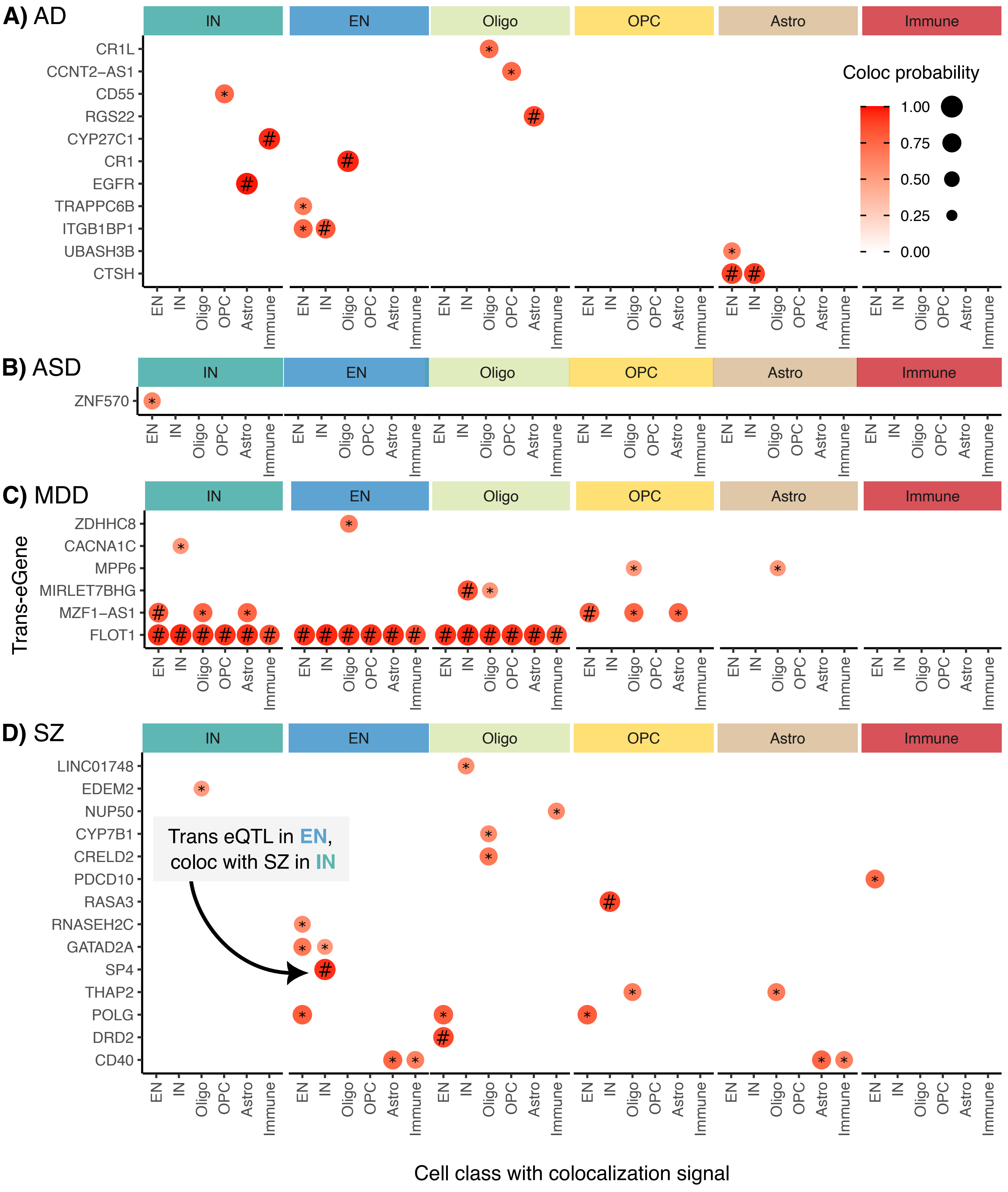


**Figure S15: Genes with a trans eQTL and a disease colocalization signal.** Genes with a trans eQTL (shown in inset of each cell class) that have a disease colocalization signal (shown on x-axis). For example, the gene SP4 has a trans eQTL detected in EN and regulatory variants for this gene colocalize with SZ risk in IN. Color and size of circle indicates posterior probability from colocalization analysis. Results are shown for **A**) AD and **B**) ASD **C**) MDD and **D**) SZ. '#' indicates colocalization probability $> 0.8$, and '*' indicates colocalization probability.
